# Integrating Artificial Intelligence and Precision Medicine to Characterize JAK-STAT Pathway Alterations in FOLFOX-Treated Colorectal Cancer in Disproportionately Affected Groups

**DOI:** 10.1101/2025.11.27.25341180

**Authors:** Fernando C. Diaz, Brigette Waldrup, Francisco G. Carranza, Sophia Manjarrez, Enrique Velazquez-Villarreal

## Abstract

Early-onset colorectal cancer (EOCRC) continues to rise, with the steepest increases observed among Hispanic/Latino (H/L) populations, underscoring the urgency of identifying ancestry- and treatment-specific biomarkers. The JAK-STAT signaling axis plays a central role in colorectal tumor biology, yet its relevance under FOLFOX-based chemotherapy in EOCRC remains poorly defined. In this study, we evaluated 2,515 colorectal cancer (CRC) cases (266 H/L; 2,249 non-Hispanic White [NHW]), stratifying analyses by ancestry, age of onset, and FOLFOX exposure. Statistical comparisons were performed using Fisher’s exact and chi-square tests, and survival patterns were assessed via Kaplan-Meier analysis. To extend conventional analytics, we deployed AI-HOPE and AI-HOPE-JAK-STAT, conversational artificial intelligence platforms capable of harmonizing genomic, clinical, demographic, and treatment variables through natural language queries, to accelerate multi-parameter biomarker exploration. JAK-STAT pathway alterations showed marked variation by ancestry and treatment context. Among H/L EOCRC cases, alterations were significantly enriched in patients who did not receive FOLFOX compared with those who did (21.2% vs. 4.1%; p = 0.003). A similar pattern emerged in late-onset CRC (LOCRC) NHW patients, where alterations were more frequent without FOLFOX exposure (13.3% vs. 7.5%; p = 0.0002). Notably, JAK-STAT alterations were significantly more common in untreated H/L EOCRC compared with untreated NHW EOCRC (21.2% vs. 9.9%; p = 0.002). Survival analyses revealed that JAK-STAT pathway alterations conferred improved overall survival across several NHW strata, including EOCRC treated with FOLFOX (p = 0.0008), EOCRC not treated with FOLFOX (p = 0.07), and LOCRC not treated with FOLFOX (p = 0.01). These findings suggest that JAK-STAT alterations may function as ancestry- and treatment-dependent prognostic markers in EOCRC, particularly among disproportionately affected H/L patients. The integration of AI-enabled platforms streamlined analyses and reveals the potential of artificial intelligence to accelerate discovery and advance precision medicine for populations historically underrepresented in cancer genomics research.

## 1. Introduction

Colorectal cancer (CRC) remains a major global health burden, ranking among the leading causes of cancer incidence and mortality according to worldwide estimates (1–3). While overall CRC rates in older adults have stabilized or declined due to improved screening and prevention, early-onset colorectal cancer (EOCRC)-diagnosed before age 50, has risen sharply across multiple regions, including the United States, Europe, Australia, and parts of Latin America (3–6). Projections indicate that by 2030, EOCRC may become the leading cause of cancer-related deaths among individuals aged 20-49 (4). This epidemiologic shift is especially concerning for Hispanic/Latino (H/L) populations, who have experienced some of the steepest increases in EOCRC incidence and poorer outcomes relative to non-Hispanic White (NHW) populations (7, 12). Recent multi-omics efforts, including those led by our group, have begun highlighting ancestry-specific molecular features in H/L EOCRC, revealing distinct alterations across canonical cancer pathways such as MAPK, PI3K, WNT, TGF-β, and JAK-STAT (1, 8–11).

Growing evidence suggests that EOCRC is biologically distinct from late-onset CRC (LOCRC), exhibiting differences in tumor mutation burden, immune microenvironment, methylation patterns, and pathway-level disruptions (14–19). Yet most genomic and clinical studies have disproportionately sampled NHW patients, leaving key gaps in understanding the molecular drivers of EOCRC within H/L communities (20). Addressing this gap is essential for developing precision oncology strategies that are equitable and responsive to population-specific needs.

Among the pathways implicated in CRC progression, the JAK-STAT signaling axis plays an essential regulatory role in inflammation-driven tumorigenesis, immune evasion, cellular differentiation, and epithelial-mesenchymal transition (21–26). Dysregulation of JAK-STAT components-including STAT3, JAK1/2, and upstream cytokines such as IL-6-has been linked to CRC aggressiveness, metastatic potential, and resistance to chemotherapy and targeted agents (27–29). Recent studies further highlight the pathway’s role in modulating response to 5-fluorouracil (5-FU) and oxaliplatin, the backbone agents in FOLFOX therapy (19, 30). For example, Xu et al. (19) demonstrated that TIMP-2 promotes 5-FU resistance through JAK-STAT activation, while Pennel et al. (30) identified JAK-STAT3 as a potential therapeutic target in stromal-rich CRC subtypes. Additional mechanistic studies also show that lncRNAs such as FAM30A and AB073614 exert tumor-suppressive or pro-metastatic functions via modulation of JAK-STAT activity (20, 22, 32). Despite these advances, the prognostic relevance of JAK-STAT alterations in the specific context of FOLFOX-treated EOCRC, especially among disproportionately affected H/L patients, remains virtually unexplored.

FOLFOX (folinic acid, fluorouracil, and oxaliplatin) is the standard first-line regimen for metastatic microsatellite-stable (MSS) CRC lacking actionable mutations (30). However, increasing evidence suggests that EOCRC patients may experience higher toxicity, poorer survival, and distinct mutational consequences following FOLFOX exposure compared to LOCRC patients (14, 32). Understanding whether ancestry-specific JAK-STAT pathway alterations influence FOLFOX responsiveness is therefore essential for guiding therapeutic decisions in diverse EOCRC populations.

Advances in artificial intelligence (AI) now offer powerful opportunities to interrogate these complex biological relationships. Large language models and conversational AI systems are increasingly being used to integrate clinical, genomic, imaging, and molecular data, enabling dynamic and interpretable precision oncology analyses (33–38). Our AI-HOPE platform, and its specialized module, AI-HOPE-JAK-STAT, represents a next-generation approach to harmonizing multi-omics datasets, conducting pathway-level comparisons, and identifying prognostic biomarkers through natural-language-driven analytic workflows (1). These systems address long-standing challenges in traditional bioinformatics pipelines, such as manual data curation, rigid query structures, and limited scalability (34–37).

The JAK-STAT signaling axis has emerged as a clinically relevant determinant of chemotherapy response in CRC, particularly in the context of FOLFOX-based cytotoxic regimens. Activation of STAT3 and upstream cytokines such as IL-6 promotes tumor cell survival, stromal remodeling, and resistance to 5-fluorouracil (5-FU) and oxaliplatin through modulation of EMT, DNA repair pathways, and anti-apoptotic programs (19, 21, 26, 30). Recent mechanistic studies have shown that TIMP-2 and other extracellular matrix regulators enhance 5-FU resistance through JAK-STAT activation (19), while STAT3 hyperactivation, driven by IL-6 trans-signaling or lncRNA dysregulation, confers reduced sensitivity to oxaliplatin and fosters more aggressive tumor phenotypes (20, 22, 29, 39–43). Clinically, tumors with high STAT3 activity display poorer responses to FOLFOX, increased recurrence rates, and a more immunosuppressive tumor microenvironment characterized by M2 macrophage polarization and reduced CD8⁺ T-cell infiltration (24, 25, 44). Conversely, preclinical inhibition of JAK1/2 or STAT3 has been shown to restore chemosensitivity to 5-FU and oxaliplatin, supporting the pathway as a potential therapeutic target for chemoresistant CRC (30, 45). These findings underscore the need to evaluate JAK-STAT alterations not only as molecular markers of disease biology but also as potential predictors of FOLFOX (46, 47) responsiveness, with implications for tailoring treatment intensity, considering JAK/STAT-directed agents, and improving outcomes in high-risk EOCRC populations.

Here, we integrate conventional statistical analyses with AI-HOPE-enabled exploration to characterize JAK-STAT pathway alterations across 2,515 CRC cases stratified by ancestry, age of onset, and FOLFOX treatment. Focusing specifically on disproportionately affected H/L EOCRC patients, we examine how JAK-STAT alterations relate to mutation prevalence patterns and overall survival outcomes. This combined computational and AI-driven framework establishes a foundation for developing ancestry-informed, treatment-responsive biomarkers and demonstrates how AI can accelerate translational precision oncology for underserved populations.

## 2. Materials and Methods

### 2.1 Data sources, cohort construction, and case selection

We performed a retrospective multi-cohort analysis using de-identified clinical and genomic data from publicly accessible CRC datasets available via cBioPortal, including TCGA Colorectal Adenocarcinoma (PanCancer Atlas), MSK-CHORD 2024, and the AACR GENIE Biopharma Collaborative (BPC) CRC dataset. These repositories were selected because they include harmonized somatic variant calls, treatment metadata, and patient-level demographics suitable for ancestry- and therapy-specific analyses. Only individuals with histologically confirmed colon, rectal, or colorectal adenocarcinoma and available next-generation sequencing data from the primary tumor were eligible. When multiple tumor samples were available for a single patient, one record was chosen at random to avoid duplicate representation.

### 2.2 Classification of disproportionately affected populations

Ancestry categories were derived primarily from self-reported ethnicity labels within the original datasets. Patients were categorized as H/L if annotated as “Hispanic,” “Latino,” “Spanish origin,” or similar descriptors. When ethnicity fields were absent, validated surname-based algorithms were applied to identify probable H/L ancestry. The comparison group consisted of NHW individuals meeting equivalent inclusion criteria. EOCRC was defined as diagnosis before age 50, whereas LOCRC was defined as diagnosis at or after age 50.

### 2.3 Identification of FOLFOX-treated and non-FOLFOX groups

Treatment information was curated from structured therapeutic annotations and free-text medication fields. Patients were assigned to the FOLFOX group if documentation confirmed exposure to all three components, 5-fluorouracil (5-FU), leucovorin, and oxaliplatin, administered concurrently or sequentially within the same line of therapy. Regimen timing and overlap were reviewed to ensure concordance with standard-of-care FOLFOX administration for microsatellite-stable disease. Individuals who did not receive all three agents were classified as non-FOLFOX.

### 2.4 JAK-STAT pathway gene set compilation and molecular alteration definition

A comprehensive JAK-STAT gene list was assembled through integration of pathway databases (KEGG, Reactome), prior CRC pathway-focused publications, and recent mechanistic studies linking JAK-STAT dysregulation to chemotherapy resistance, tumor progression, or immune modulation (19–30, 43–45). Mutation data were extracted from cBioPortal’s MAF/variant tables and filtered to retain non-synonymous coding alterations, including missense, nonsense, frameshift, splice-site, and start-loss variants. Pathway alteration status was defined as the presence of ≥1 qualifying mutation in any JAK-STAT pathway gene.

### 2.5 Statistical analysis

Differences in pathway alteration frequencies between ancestry groups, age categories, and treatment-status groups were evaluated using Fisher’s exact tests or chi-square tests. Continuous variables were compared using the Mann-Whitney U test. Overall survival (OS) was defined as time from diagnosis or sequencing (depending on dataset availability) until death or last follow-up. Kaplan-Meier curves were generated for all major subgroups, and survival differences were assessed using the log-rank test. Cox proportional hazards models were used to estimate hazard ratios (HRs) with 95% confidence intervals (CIs). Multivariable analyses adjusted for age, sex, tumor site, MSI status, and treatment group. All analyses were conducted in R (v4.3.2), with two-sided p-values <0.05 considered statistically significant.

### 2.6 AI-HOPE-enabled data harmonization, pathway interrogation, and analytic refinement

To accelerate dataset harmonization and improve analytic rigor, we used the AI-HOPE and AI-HOPE-JAK-STAT (2) conversational artificial intelligence systems to perform structured, natural-language-driven exploration prior to formal statistical testing. These agents integrate clinical variables, genomic alterations, ancestry metadata, and treatment histories into a unified analytic layer capable of automated filtering and dynamic sub-cohort construction.

AI-HOPE-JAK-STAT was used to identify and verify patients meeting combined clinical and molecular criteria, including ancestry, EOCRC/LOCRC status, FOLFOX exposure, and JAK-STAT alteration status. Generate rapid pathway-specific mutation frequency tables with stratification by ancestry and treatment group. Conduct iterative survival-driven subgroup scans, enabling rapid detection of candidate prognostic signals for downstream validation. Perform natural-language queries. This AI-assisted approach reduced manual curation burden, minimized transcription errors, and enabled systematic cross-comparison across multiple datasets. All AI-generated subgroup identifications and frequency patterns were manually verified and subsequently tested using standard statistical methods to ensure reproducibility and analytic transparency.

## 3. Results

### 3.1 Clinical and Demographic Characteristics of H/L and NHW Cohorts

Table 1 summarizes the major clinical and demographic attributes of the H/L and NHW CRC cohorts evaluated in this study. Among the 2,515 total cases, 266 (10.6%) were identified as H/L and 2,249 (89.4%) as NHW. Patterns of age at disease onset and FOLFOX exposure differed noticeably between groups. H/L patients exhibited a higher representation of EOCRC receiving FOLFOX (27.4%) compared with NHW patients (16.7%). Similarly, EOCRC cases not exposed to FOLFOX were proportionally more frequent in the H/L cohort (19.5%) than in the NHW cohort (13.4%). In contrast, LOCRC treated with FOLFOX was more common among NHW patients (40.9%) compared with H/L patients (34.2%).

**Table 1.** Overview of key clinical and demographic features for Hispanic/Latino (H/L) and Non-Hispanic White (NHW) patients with CRC, including distributions by diagnostic age, receipt of FOLFOX therapy, tumor classification, and detailed ethnicity categories.

| Clinical Feature | H/L Cohort n (%) | NHW Cohort n (%) | Total (n) |
| --- | --- | --- | --- |
| **Age at Disease Onset** |  |  |  |
| EOCRC Treated with FOLFOX | 73 (27.4%) | 375 (16.7%) | 448 |
| EOCRC Not Treated with FOLFOX | 52 (19.5%) | 302 (13.4%) | 354 |
| LOCRC Treated with FOLFOX | 91 (34.2%) | 919 (40.9%) | 1010 |
| LOCRC Not Treated with FOLFOX | 50 (18.8%) | 653 (29.0%) | 703 |
| **Sex** |  |  |  |
| Male | 158 (59.4%) | 1267 (56.3%) | 1425 |
| Female | 108 (40.6%) | 982 (43.7%) | 1090 |
| **Sample type** |  |  |  |
| Primary Tumor | 266 (100.0%) | 2249 (100.0%) | 2515 |
| **State at Diagnosis** |  |  |  |
| Stage 1-3 | 156 (58.6%) | 1236 (55.0%) | 1392 |
| Stage 4 | 108 (40.6%) | 1005 (44.7%) | 1113 |
| NA | 2 (0.8%) | 8 (0.4%) | 10 |
| **Cancer Type** |  |  |  |
| Colon Adenocarcinoma | 164 (61.7%) | 1328 (59.0%) | 1492 |
| Rectal Adenocarcinoma | 64 (24.1%) | 646 (28.7%) | 710 |
| Colorectal Adenocarcinoma | 38 (14.3%) | 275 (12.2%) | 313 |
| **Ethnicity** |  |  |  |
| Spanish NOS / Hispanic NOS / Latino NOS | 230 (86.5%) | 0 (0.0%) | 230 |
| Mexican (includes Chicano) | 30 (11.3%) | 0 (0.0%) | 30 |
| Hispanic Category 2 | 2 (0.8%) | 0 (0.0%) | 2 |
| Hispanic Category 1 | 1 (0.4%) | 0 (0.0%) | 1 |
| Other Spanish / Hispanic | 3 (1.1%) | 0 (0.0%) | 3 |
| Not Hispanic/Latino | 0 (0.0%) | 2249 (100.0%) | 2249 |

Sex distribution was comparable between populations, with males comprising 59.4% of H/L cases and 56.3% of NHW cases. All tumors represented in both cohorts were primary malignancies. Disease stage at diagnosis generally followed similar trends, with a slightly higher percentage of NHW patients presenting with metastatic (stage 4) disease (44.7%) relative to H/L patients (40.6%).

Tumor site distributions were also broadly aligned across groups. Colon adenocarcinoma was the most common diagnosis in both H/L (61.7%) and NHW (59.0%) patients, followed by rectal adenocarcinoma and combined colorectal classifications.

Ethnicity annotations showed complete separation between groups, reflecting the curated cohort definitions. Among H/L patients, the majority were categorized as “Spanish NOS / Hispanic NOS / Latino NOS” (86.5%), with smaller fractions identified as Mexican/Chicano (11.3%) or other Hispanic subcategories. As expected, all NHW patients were classified as not H/L.

### 3.2 Genomic Comparisons Across Age Groups and Ancestral Backgrounds

#### 3.2.1 Baseline Characteristics of the H/L and NHW Cohorts

Analysis of the clinical profiles demonstrated notable differences in the distribution of age at onset and FOLFOX exposure between H/L and NHW CRC patients. EOCRC treated with FOLFOX represented a substantially larger share of the H/L cohort (27.4%) compared to NHW patients (16.7%), and EOCRC cases without FOLFOX therapy were also proportionally higher in H/L individuals (19.5% vs. 13.4%). In contrast, LOCRC treated with FOLFOX occurred more frequently among NHW patients (40.9%) relative to H/L patients (34.2%), reflecting differences in treatment patterns or age-associated therapeutic decision-making across populations.

Sex distribution was similar between groups, with males comprising the majority of cases in both cohorts. All tumors analyzed were derived from primary tumor specimens. Disease stage at diagnosis showed modest variation: a slightly greater proportion of NHW patients presented with stage 4 disease (44.7%) compared to H/L patients (40.6%), whereas stages 1-3 were more common in the H/L group (58.6% vs. 55.0%).

Tumor location patterns were broadly consistent between groups, with colon adenocarcinoma being the predominant subtype in both H/L (61.7%) and NHW (59.0%) patients, followed by rectal adenocarcinoma and combined colorectal classifications. As anticipated based on cohort definitions, ethnicity categories demonstrated complete separation: all NHW cases were classified as non-Hispanic/non-Latino, whereas the H/L cohort predominantly comprised individuals annotated as “Spanish NOS / Hispanic NOS / Latino NOS,” with additional representation of Mexican/Chicano and smaller Hispanic subcategories.

#### 3.2.2 Clinical Distribution of NHW Patients by Age at Onset, Treatment Status, and Tumor Characteristics

Table 2b summarizes the distribution of EOCRC and LOCRC cases among NHW patients in relation to FOLFOX therapy, clinical features, and tumor subtypes. Within the NHW cohort, EOCRC represented a sizeable proportion of cases, with 16.7% receiving FOLFOX and 13.4% not receiving FOLFOX. In contrast, LOCRC constituted the majority, and these patients showed a higher prevalence of FOLFOX exposure (40.9%) compared with their non-treated counterparts (29.0%). These patterns indicate that, among NHW individuals, age at diagnosis is strongly associated with likelihood of receiving FOLFOX, with treatment being more commonly administered in older adults.

**Table 2.** Comparative clinical and genomic profiles of early-onset and late-onset colorectal cancer (CRC) patient cohorts. This table outlines clinical and molecular distinctions, including JAK-STAT pathway alterations and mutation burden, across key subgroups: (a) Early-Onset CRC (EOCRC) versus Late-Onset CRC (LOCRC) within Hispanic/Latino (H/L) patients; (b) EOCRC versus LOCRC within Non-Hispanic White (NHW) patients; (c) EOCRC comparisons between H/L and NHW cohorts; and (d) EOCRC versus LOCRC comparisons between H/L treated and not treated with FOLFOX. Comparisons include median age at diagnosis, total mutation counts, and the prevalence of selected JAK-STAT pathway gene alterations, stratified by both ethnicity and age category.

| Clinical Feature | Early-Onset Hispanic/Latino<br>Treated with FOLFOX<br>n (%) | Early-Onset Hispanic/Latino<br>Not Treated with FOLFOX<br>n (%) | p-value | Late-Onset Hispanic/Latino<br>Treated with FOLFOX<br>n (%) | Late-Onset Hispanic/Latino<br>Not Treated with FOLFOX<br>n (%) | p-value |
| --- | --- | --- | --- | --- | --- | --- |
| Median Diagnosis Age (IQR) | 42 (36-47) | 40 (34-43) | 0.05411 | 59 (54-66) | 62 (56-70) | **0.04865** |
| Median Mutation Count | 7 (5-8) | 7 (5-20) | 0.09735 | 8 (6-9) [NA=1] | 7 (5.25-9) | 0.6507 |
| Median TMB (IQR) | 6.3 (4.5-7.8) [NA=15] | 5.5 (3.4-8.3) [NA=2] | 0.1719 | 6.1 (4.9-7.8) [NA=10] | 6.9 (5.6-9.0) [NA=2] | 0.04389 |
| Median FGA | 0.18 (0.03-0.27) [NA=6] | 0.19 (0.03-0.29) | 0.7661 | 0.15 (0.06-0.25) [NA=7] | 0.21 (0.04-0.3) [NA=2] | 0.5464 |
| **STAT5B Mutation** |  |  |  |  |  |  |
| Present | 0 (0.0%) | 5 (9.6%) | **0.01108** | 1 (1.1%) | 1 (1.1%) | 1 |
| Absent | 73 (100.0%) | 47 (90.4%) |  | 90 (98.9%) | 90 (98.9%) |  |

(b)
| Clinical Feature | Early-Onset NHW<br>Treated with FOLFOX<br>n (%) | Early-Onset NHW<br>Not Treated with FOLFOX<br>n (%) | p-value | Late-Onset NHW<br>Treated with FOLFOX<br>n (%) | Late-Onset NHW<br>Not Treated with FOLFOX<br>n (%) | p-value |
| --- | --- | --- | --- | --- | --- | --- |
| Median Diagnosis Age (IQR) | 43 (37-48) | 44 (38-47) | 0.5646 | 63 (57-69) | 66 (57-74) | **4.146E-07** |
| Median Mutation Count | 6 (5-8) [NA=4] | 7 (5-9) [NA=2] | 0.1258 | 7 (5-9) [NA=10] | 8 (6-12) [NA=3] | **1.22E-05** |
| Median TMB (IQR) | 5.7 (4.1-6.9) | 5.7 (4.1-7.8) | 0.4214 | 6.1 (4.3-8.2) | 6.6 (4.9-10.4) | **0.0002854** |
| Median FGA | 0.14 (0.04-0.24) [NA=4] | 0.15 (0.04-0.23) [NA=2] | 0.5589 | 0.16 (0.06-0.28) [NA=6] | 0.15 (0.05-0.27) [NA=5] | 0.1929 |
| **JAK3 Mutation** |  |  |  |  |  |  |
| Present | 4 (1.1%) | 14 (4.6%) | **0.006502** | 14 (2.6%) | 28 (4.3%) | 0.9409 |
| Absent | 371 (98.9%) | 288 (95.4%) |  | 288 (97.4%) | 625 (95.7%) |  |

(c)
| Clinical Feature | Early-Onset Hispanic/Latino<br>Treated with FOLFOX<br>n (%) | Early-Onset NHW<br>Treated with FOLFOX<br>n (%) | p-value | Early-Onset Hispanic/Latino<br>Not Treated with FOLFOX<br>n (%) | Early-Onset NHW<br>Not Treated with FOLFOX<br>n (%) | p-value |
| --- | --- | --- | --- | --- | --- | --- |
| Median Diagnosis Age (IQR) | 42 (36-47) | 43 (37-48) | 0.08467 | 40 (34-43) | 44 (38-47) | **0.0006016** |
| Median Mutation Count | 7 (5-8) | 6 (5-8) [NA=4] | 0.942 | 7 (5-20) | 7 (5-9) [NA=2] | 0.2601 |
| Median TMB (IQR) | 6.3 (4.5-7.8) [NA=15] | 5.7 (4.1-6.9) | 0.05806 | 5.5 (3.4-8.3) [NA=2] | 5.7 (4.1-7.8) | 0.5732 |
| Median FGA | 0.18 (0.03-0.27) [NA=6] | 0.14 (0.04-0.24) [NA=4] | 0.5556 | 0.19 (0.03-0.29) | 0.15 (0.04-0.23) [NA=2] | 0.3612 |
| **STAT5B Mutation** |  |  |  |  |  |  |
| Present | 0 (0.0%) | 5 (1.3%) | 1 | 5 (9.6%) | 7 (2.3%) | **0.01994** |
| Absent | 73 (100.0%) | 370 (98.7%) |  | 47 (90.4%) | 295 (97.7%) |  |

Sex distribution did not differ markedly across groups, as males comprised just over half of NHW cases (56.3%), with females representing 43.7%. All NHW tumor samples were derived from primary tumors, ensuring consistency across molecular and clinical analyses. With respect to disease stage, more than half of NHW patients were diagnosed with non-metastatic (stage 1-3) disease (55.0%), while 44.7% presented with stage 4 disease, reflecting the substantial burden of advanced tumors in this group.

Tumor classification patterns were also consistent with population-level CRC distributions: colon adenocarcinoma was the predominant diagnosis (59.0%), followed by rectal adenocarcinoma (28.7%) and combined colorectal adenocarcinoma (12.2%). Ethnicity categories confirmed the cohort’s intended demographic composition, with all NHW patients recorded as non-Hispanic/non-Latino.

#### 3.2.3 Ethnic Patterns Among EOCRC Cases by Treatment Status

Table 2C highlights key differences in the distribution of EOCRC cases between H/L and NHW patients across treatment groups. Among individuals receiving FOLFOX, EOCRC represented a substantially larger proportion of the H/L cohort (27.4%) compared with NHW patients (16.7%), indicating a higher relative burden of treated EOCRC in H/L populations. A similar pattern emerged in the non-FOLFOX EOCRC group, where H/L patients again showed greater representation (19.5% vs. 13.4% in NHW patients). These differences were observed despite similar sex distributions between groups and the exclusive use of primary tumor specimens across all analyses.

While tumor stage distributions were broadly comparable, H/L patients presented with slightly more stage 1-3 disease (58.6%) than NHW patients (55.0%), whereas stage 4 diagnoses were marginally more common in NHW individuals (44.7% vs. 40.6%). Tumor type patterns also showed consistent trends across ethnicities: colon adenocarcinoma was the predominant diagnosis in both groups, though rectal adenocarcinoma was somewhat more frequent among NHW patients. As expected, ethnicity categorizations reflected distinct population definitions, with all NHW patients classified as non-Hispanic/non-Latino and the H/L cohort comprising several Hispanic subgroups, the majority of whom were annotated as “Spanish NOS / Hispanic NOS / Latino NOS.”

### 3.3 JAK-STAT Pathway Alterations by Age, Ancestry, and Treatment Status

A detailed assessment of JAK-STAT pathway alterations across age-of-onset categories, ancestry groups, and FOLFOX exposure is presented in Tables 3a-3d. Distinct patterns emerged within and between populations, highlighting context-specific associations between pathway alterations and treatment status.

**Table 3.** JAK-STAT Alteration Frequencies by Age, Ancestry, and FOLFOX Status in CRC. The table is divided into four comparative panels: **(3a)** EOCRC versus LOCRC H/L patients separated by treatment status; **(3b)** analogous comparisons within NHW EOCRC and LOCRC groups; **(3c)** cross-ancestry evaluation of EOCRC H/L and NHW patients stratified by FOLFOX receipt; and **(3d)** the same ancestry-based comparison in LOCRC. Mutation data encompass the primary genes governing JAK-STAT signaling. Significant differences identified by Chi-square or Fisher’s exact testing (p < 0.05) are marked accordingly. Collectively, these stratified analyses illustrate how age, ancestral background, and chemotherapy exposure intersect with variations in JAK-STAT pathway disruption in CRC.

| Pathway Alterations | Early-Onset Hispanic/Latino<br>Treated with FOLFOX<br>n (%) | Early-Onset Hispanic/Latino<br>Not Treated with FOLFOX<br>n (%) | p-value | Late-Onset Hispanic/Latino<br>Treated with FOLFOX<br>n (%) | Late-Onset Hispanic/Latino<br>Not Treated with FOLFOX<br>n (%) | p-value |
| --- | --- | --- | --- | --- | --- | --- |
| JAK/STAT Alterations Present | 3 (4.1%) | 11 (21.2%) | 0.003851 | 10 (11.0%) | 3 (6.0%) | 0.3811 |
| JAK/STAT Alterations Absent | 70 (95.9%) | 41 (78.8%) |  | 81 (89.0%) | 47 (94.0%) |  |

(b)
| Pathway Alterations | Early-Onset NHW<br>Treated with FOLFOX<br>n (%) | Early-Onset NHW<br>Not Treated with FOLFOX<br>n (%) | p-value | Late-Onset NHW<br>Treated with FOLFOX<br>n (%) | Late-Onset NHW<br>Not Treated with FOLFOX<br>n (%) | p-value |
| --- | --- | --- | --- | --- | --- | --- |
| JAK/STAT Alterations Present | 27 (7.2%) | 30 (9.9%) | 0.9979 | 69 (7.5%) | 87 (13.3%) | 0.0002036 |
| JAK/STAT Alterations Absent | 348 (92.8%) | 372 (123.2%) |  | 850 (92.5%) | 566 (86.7%) |  |

(c)
| Pathway Alterations | Early-Onset Hispanic/Latino<br>Treated with FOLFOX<br>n (%) | Early-Onset NHW<br>Treated with FOLFOX<br>n (%) | p-value | Early-Onset Hispanic/Latino<br>Not Treated with FOLFOX<br>n (%) | Early-Onset NHW<br>Not Treated with FOLFOX<br>n (%) | p-value |
| --- | --- | --- | --- | --- | --- | --- |
| JAK/STAT Alterations Present | 3 (4.1%) | 27 (7.2%) | 0.4472 | 11 (21.2%) | 30 (9.9%) | 0.002843 |
| JAK/STAT Alterations Absent | 70 (95.9%) | 348 (92.8%) |  | 41 (78.8%) | 372 (123.2%) |  |

(d)
| Pathway Alterations | Late-Onset Hispanic/Latino<br>Treated with FOLFOX<br>n (%) | Late-Onset NHW<br>Treated with FOLFOX<br>n (%) | p-value | Late-Onset Hispanic/Latino<br>Not Treated with FOLFOX<br>n (%) | Late-Onset NHW<br>Not Treated with FOLFOX<br>n (%) | p-value |
| --- | --- | --- | --- | --- | --- | --- |
| JAK/STAT Alterations Present | 10 (11.0%) | 69 (7.5%) | 0.3296 | 3 (6.0%) | 87 (13.3%) | 0.1857 |
| JAK/STAT Alterations Absent | 81 (89.0%) | 850 (92.5%) |  | 47 (94.0%) | 566 (86.7%) |  |

#### 3.3.1 Within-ancestry comparisons

Among H/L patients, EOCRC cases demonstrated marked differences by treatment group: JAK-STAT alterations were detected in 21.2% of individuals who did not receive FOLFOX compared with 4.1% of FOLFOX-treated patients (p = 0.003) (Table 3a). In LOCRC H/L patients, alteration frequencies were modest and did not differ significantly between treated (11%) and untreated (6%) groups (Table 3a).

Among NHW patients, EOCRC cases showed comparable alteration rates across treatment categories, with 7.2% in the FOLFOX-treated group and 9.9% in the non-treated group (p = 0.99) (Table 3b). However, a notable divergence was observed in LOCRC NHW patients: JAK-STAT alterations were significantly less common among those receiving FOLFOX (7.5%) compared with those not treated with FOLFOX (13.3%, p = 0.0002) (Table 3b), suggesting a potential interaction between treatment exposure and age-associated tumor biology in this group.

#### 3.3.2 Between-ancestry comparisons

In EOCRC, JAK-STAT alteration frequencies were generally comparable between H/L and NHW patients receiving FOLFOX (4.1% vs. 7.2%, p = 0.44) (Table 3c). In contrast, EOCRC patients who did not receive FOLFOX showed a substantially higher prevalence of alterations in the H/L group (21.2%) relative to NHW patients (9.9%, p = 0.002) (Table 3c), indicating potential ancestry-related differences in untreated tumor biology.

Among LOCRC cases, FOLFOX-treated H/L patients exhibited a slightly higher alteration rate (11%) than NHW patients (7.5%), though the difference was not statistically significant (Table 3d). In non-FOLFOX LOCRC, H/L patients showed lower alteration frequencies than NHW patients (6% vs. 13.3%) (Table 3d), again without statistical significance. These patterns suggest that ancestry-related differences in JAK-STAT dysregulation may be most pronounced in EOCRC disease and most evident in patients who did not receive FOLFOX.

Across age, ancestry, and treatment strata, JAK-STAT pathway alterations displayed heterogeneous patterns that were not uniformly distributed across clinical subgroups. The strongest signals emerged in EOCRC, where untreated H/L patients demonstrated markedly higher alteration rates than their NHW counterparts, and in LOCRC NHW patients, where alterations were significantly less frequent in those treated with FOLFOX.

### 3.4 Frequencies of JAK-STAT Pathway Alterations Across Age, Ancestry, and Treatment Groups

Across EOCRC H/L patients (Table S1), JAK-STAT pathway mutations were generally rare, with most genes showing very low alteration frequencies. The only statistically significant difference was observed for STAT5B, which was mutated in 9.6% of non-FOLFOX patients and absent in all FOLFOX-treated individuals (p = 0.01108). Although alterations remained infrequent overall, the selective enrichment of STAT5B mutations in untreated EOCRC H/L cases suggests a potential treatment-related distinction within this subgroup.

In LOCRC H/L patients (Table S2), JAK-STAT alterations were similarly uncommon, and no significant differences were identified between FOLFOX-treated and untreated groups for any gene. Mutation frequencies remained low across all loci. Among NHW EOCRC patients (Table S3), overall alteration rates were also low; however, JAK3 mutations were significantly more prevalent in untreated cases (4.6%) than in treated cases (1.1%, p = 0.006), representing the only gene-level difference within this subgroup. In LOCRC NHW patients (Table S4), JAK-STAT mutations were infrequent and no statistically significant differences were noted between treatment groups.

Age-stratified analyses within the H/L cohort showed consistent patterns. Among FOLFOX-treated patients (Table S5), no gene exhibited significant differences between EOCRC and LOCRC disease. In contrast, among those not treated with FOLFOX (Table S6), STAT5B remained higher in EOCRC cases (9.6%) compared with LOCRC cases (1.1%), reflecting its enrichment in younger untreated H/L patients.

Similarly, age-stratified comparisons within NHW patients demonstrated stable mutation profiles. In the FOLFOX-treated NHW cohort (Table S7), mutation frequencies did not differ meaningfully by age, aside from a nominal difference in JAK3, which was slightly elevated in EOCRC cases (though the reported percentages suggest further verification may be needed). Overall, no systematic age-related shifts were observed.

Ancestry-stratified comparisons also revealed generally consistent alteration patterns. Among EOCRC patients treated with FOLFOX (Table S8), H/L and NHW cohorts showed no significant differences in mutation frequencies for any JAK-STAT gene. In non-FOLFOX EOCRC patients (Table S9), most genes displayed similar prevalence across ancestries, though STAT5B mutations were significantly more common in H/L patients (9.6%) than NHW patients (2.3%, p = 0.01).

In LOCRC patients treated with FOLFOX (Table S10), mutation distributions were comparable between ancestries, with no statistically significant differences detected. The same pattern was observed in LOCRC non-FOLFOX patients (Table S11).

### 3.5 Mutational Landscape of the JAK-STAT Pathway

#### 3.5.1 Early-Onset H/L CRC

Figure 1a illustrates the somatic mutation profile of the JAK-STAT signaling pathway in EOCRC H/L patients (n = 122), integrating mutation class, tumor mutational burden (TMB), and FOLFOX treatment status. Overall, 14 patients (11.48%) exhibited at least one alteration affecting pathway genes. Among mutated genes, JAK1 and STAT5B were the most frequently altered (4%), followed by JAK3 (3%), STAT3 (2%), and STAT5A (1%). All remaining pathway components, including SOCS1, STAT1/4/6, JAK2, PIAS family members, and PTPRC, showed no detectable alterations in this cohort. Mutations were predominantly missense variants (green), with fewer truncating events such as frame-shift deletions (light blue), frame-shift insertions (dark blue), nonsense mutations (red), and occasional splice-site alterations (orange). Multi-hit events (black) were rare and limited to isolated cases. TMB values spanned a broad range, with a subset of hypermutated tumors represented by elevated TMB bars above the mutation matrix. These high-TMB samples did not cluster around any specific JAK-STAT gene, suggesting that elevated mutational load reflects broader tumor biology rather than pathway-specific genomic disruption. FOLFOX treatment status, shown in the annotation track (blue = treated; red = not treated), was distributed across both mutated and non-mutated samples without a discernible pattern of enrichment. This indicates that, within EOCRC H/L patients, JAK-STAT pathway alterations do not segregate by chemotherapy exposure and remain relatively infrequent across the population.

**Figure 1.**
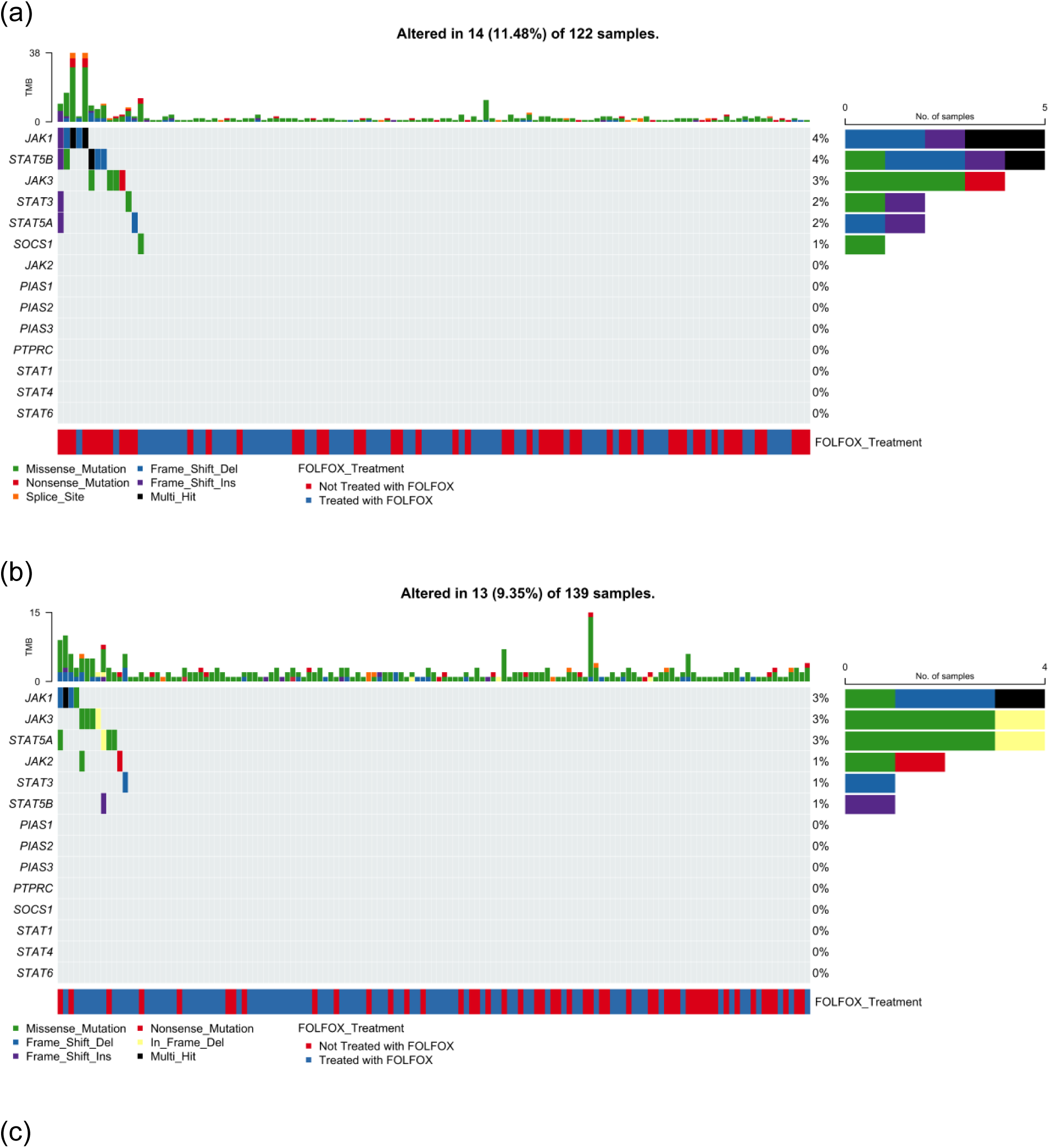
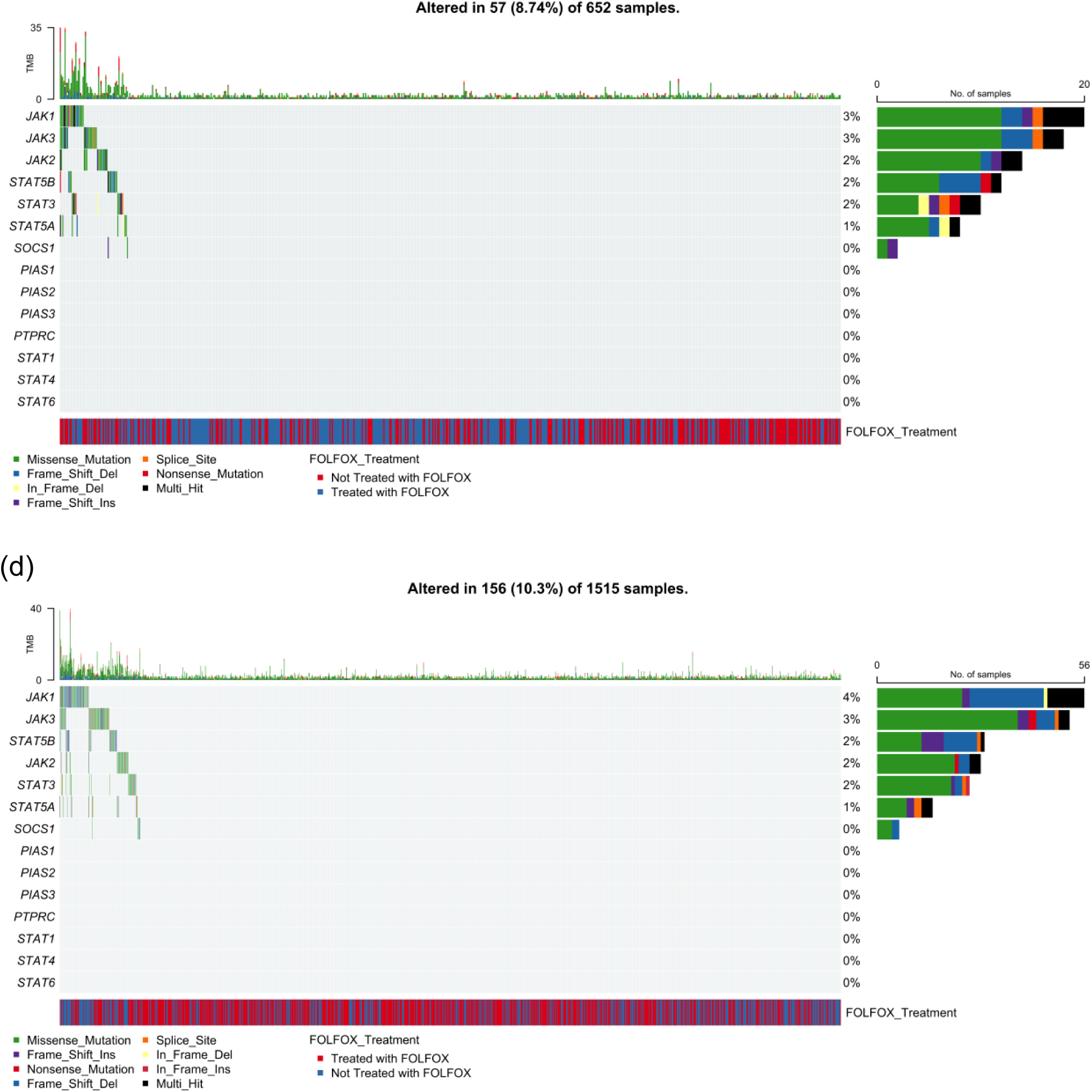
Somatic alteration patterns in JAK-STAT pathway genes among colorectal cancer (CRC) patients stratified by ancestry and diagnostic age. This figure presents oncoplots illustrating the distribution of mutations affecting major components of the JAK-STAT signaling pathway in CRC. Mutation profiles are shown separately for early- (EOCRC) and late-onset (LOCRC) disease and for Hispanic/Latino (H/L) and Non-Hispanic White (NHW) populations. Each panel depicts mutation classes, tumor mutational burden (TMB), and FOLFOX treatment status for: (a) 122 H/L patients diagnosed before age 50, (b) 139 H/L patients diagnosed at or after age 50, (c) 652 EOCRC NHW patients, and (d) 1,515 LOCRC NHW patients.

#### 3.5.2 Late-Onset H/L CRC

In LOCRC H/L patients (n = 139; Figure 1b), 13 tumors (9.35%) exhibited at least one alteration affecting the JAK-STAT signaling pathway. Similar to the EOCRC group, overall mutation frequencies were low, with alterations distributed across a limited set of pathway genes. JAK1, JAK3, and STAT5A were the most frequently mutated (each ∼3%), predominantly through missense variants (green), with occasional truncating mutations such as frame-shift deletions (light blue) and nonsense substitutions (red). STAT3 (1%) and STAT5B (1%) showed a mixture of missense and in-frame deletion events, while JAK2 demonstrated rare missense alterations. All remaining pathway components, including PIAS family genes, SOCS1, PTPRC, STAT1, STAT4, and STAT6, showed no detectable mutations in this cohort. TMB values were generally low across samples, with sporadic elevations that did not cluster around specific JAK-STAT gene mutations. FOLFOX treatment status (blue = treated; red = untreated) was evenly represented across mutated and non-mutated cases, indicating no clear enrichment of pathway alterations in relation to chemotherapy exposure. Collectively, the LOCRC H/L mutational profile demonstrates that JAK-STAT pathway disruption is rare in this subgroup and lacks treatment-associated or mutation-type-specific clustering.

#### 3.5.3 Early-Onset NHW CRC

In EOCRC NHW patients (n = 652; Figure 1c), 57 tumors (8.74%) exhibited at least one alteration in the JAK-STAT signaling pathway. The most frequently mutated genes were JAK1 and JAK3 (each ∼3%), with alterations dominated by missense variants (green), accompanied by occasional nonsense mutations (red), frame-shift deletions (light blue), and splice-site changes (orange). JAK2, STAT5B, and STAT3 each accounted for approximately 2% of cases, displaying a mixture of missense and truncating events, including multi-hit alterations (black) in a minority of tumors. Lower-frequency mutations (<2%) were observed in STAT5A, SOCS1, and select PIAS family members, while several downstream genes, including STAT1, STAT4, STAT6, and PTPRC, showed no detectable alterations. TMB varied widely across the cohort, with a notable subset of hypermutated tumors, although elevated TMB did not correspond to specific JAK-STAT gene mutations. FOLFOX treatment status (blue = treated; red = not treated) was broadly distributed, with no evidence of clustering by mutation burden or gene-specific alteration patterns. Compared with EOCRC H/L patients, the EOCRC NHW group exhibited a slightly lower overall prevalence of JAK-STAT pathway alterations, suggesting potentially distinct molecular profiles across ancestry groups.

#### 3.5.4 Late-Onset NHW CRC

In LOCRC NHW patients (n = 1,515; Figure 1d), 156 tumors (10.3%) exhibited at least one alteration in the JAK-STAT pathway. JAK1 was the most frequently mutated gene (∼4%), followed by JAK3 (∼3%) and STAT5B (∼2%), each demonstrating a diverse spectrum of mutation types dominated by missense variants (green), along with intermittent frame-shift deletions (light blue), nonsense substitutions (red), splice-site alterations (orange), and multi-hit events (black). JAK2, STAT3, and STAT5A each contributed mutations in approximately 2% of cases, with a mix of missense and truncating alterations. Lower-prevalence changes were observed in SOCS1, PIAS1, and PIAS3, whereas several pathway members, including STAT1, STAT4, STAT6, PIAS2, and PTPRC, remained unaltered in this cohort. TMB values were generally low to moderate across the population, with a distinct subset of hypermutated tumors; however, these elevated TMB cases did not cluster around specific JAK-STAT gene alterations. FOLFOX treatment status (blue = treated; red = untreated) was evenly distributed among mutated and non-mutated samples, with no clear enrichment pattern associated with chemotherapy exposure. Overall, the LOCRC NHW cohort displayed a modestly higher prevalence of JAK-STAT pathway alterations than EOCRC NHW patients, consistent with broader genomic instability observed in LOCRC, though without evidence of treatment-associated clustering.

### 3.6 Overall Survival Patterns Associated with JAK-STAT Pathway Mutations Across Clinical and Demographic Strata

To determine whether JAK-STAT pathway mutations convey prognostic significance in CRC, we performed Kaplan-Meier survival analyses within subgroups categorized by age at diagnosis, ancestral background, and receipt of FOLFOX chemotherapy.

#### 3.6.1 Early-Onset H/L Patients Treated With FOLFOX

Among EOCRC H/L patients treated with FOLFOX (Figure 2a), the presence of JAK-STAT pathway alterations did not demonstrate a meaningful association with overall survival (p = 0.68). Survival curves for altered and non-altered groups overlapped substantially throughout follow-up, indicating comparable long-term outcomes. The altered cohort showed greater variability, reflected by wider confidence intervals, largely due to the very limited number of cases carrying pathway mutations. In contrast, patients without alterations displayed a steady but gradual decline in survival after approximately 30 months, though overall survival remained relatively high across the group. These findings suggest that, within EOCRC H/L receiving FOLFOX, JAK-STAT pathway mutations do not appear to exert a significant prognostic influence, though sample size limitations in the altered subgroup warrant cautious interpretation.

**Figure 2.**
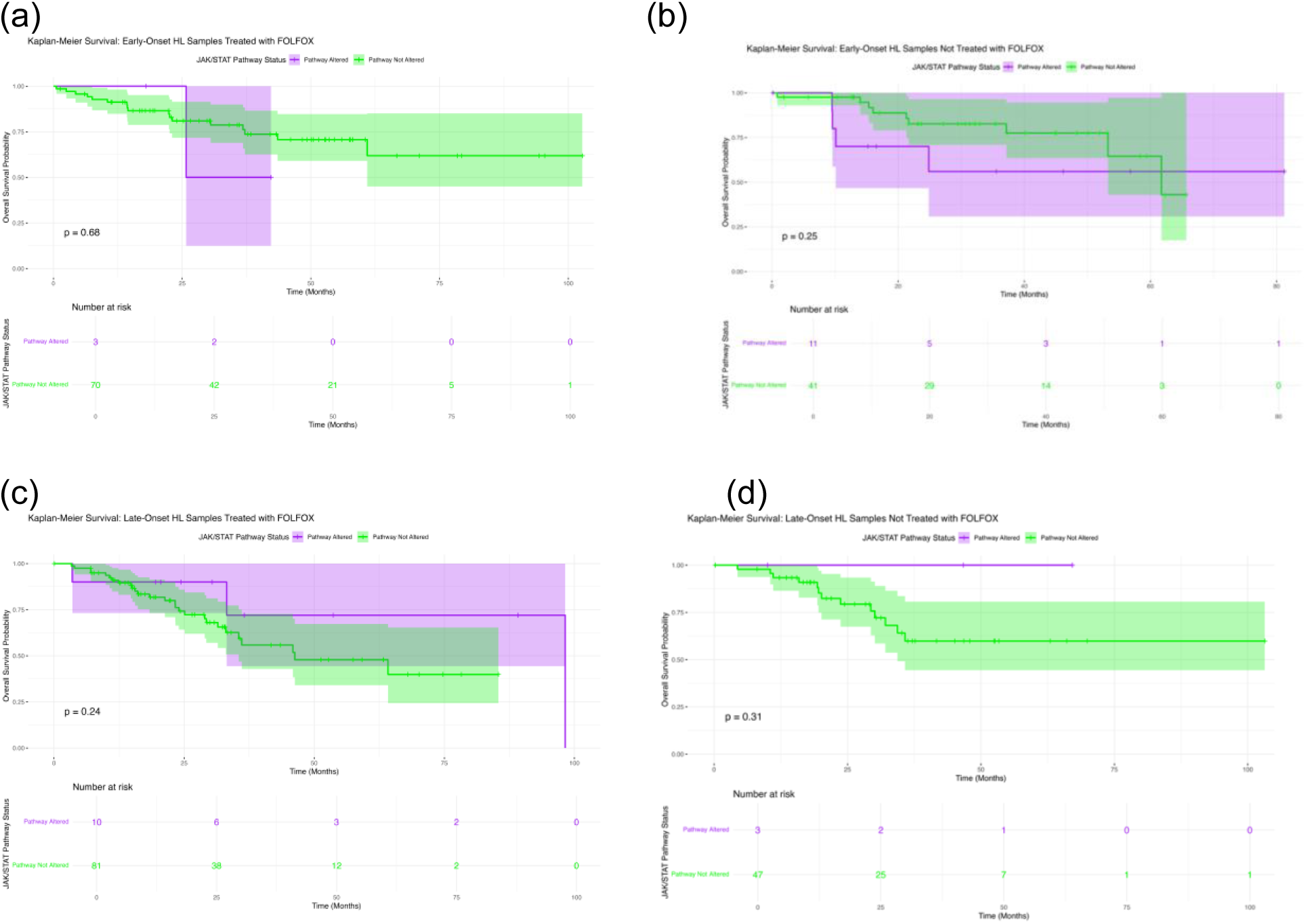
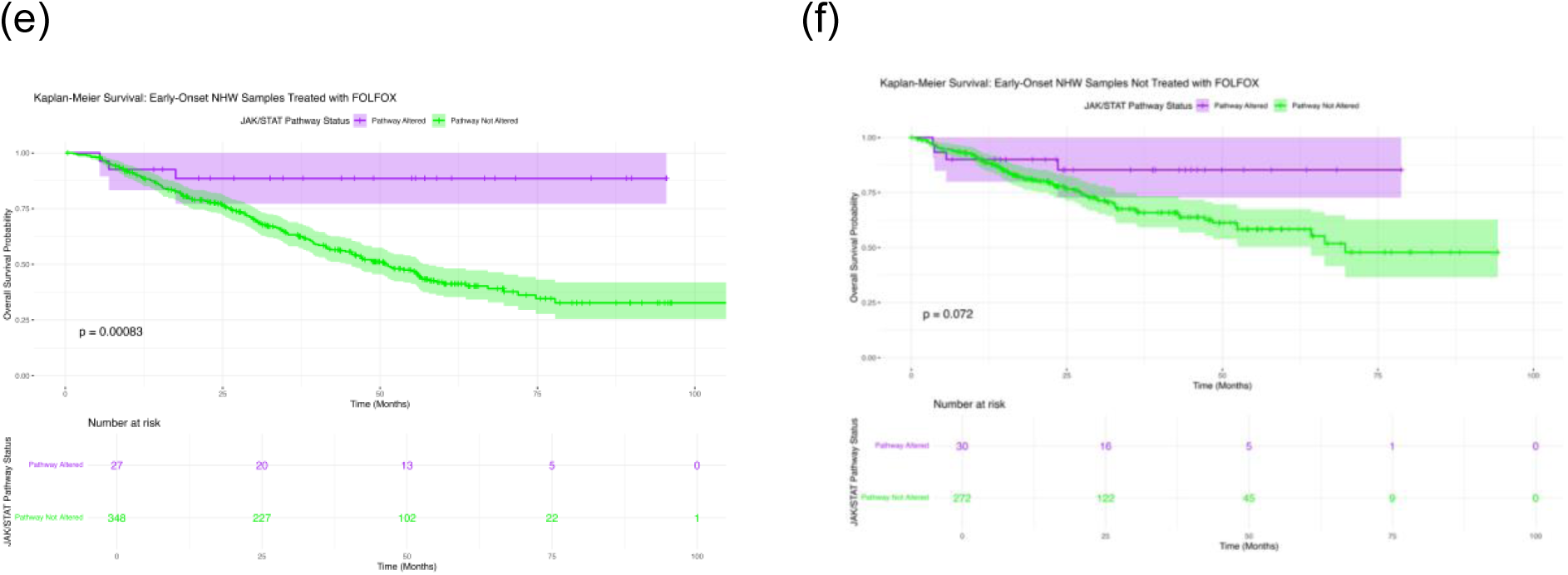
Kaplan-Meier curves illustrating overall survival stratified by JAK-STAT pathway mutation status in colorectal cancer (CRC) across age, ancestry, and treatment-defined subgroups. Survival outcomes are displayed separately for: (a) early-onset (EOCRC) Hispanic/Latino (H/L) patients who received FOLFOX, (b) EOCRC H/L patients who did not receive FOLFOX, (c) late-onset (LOCRC) H/L patients treated with FOLFOX, (d) LOCRC H/L patients not treated with FOLFOX, (e) EOCRC Non-Hispanic White (NHW) patients treated with FOLFOX, and (f) EOCRC NHW patients not treated with FOLFOX. Each panel contrasts individuals harboring JAK-STAT pathway alterations with those lacking mutations, highlighting potential survival differences within each clinical and demographic group. Confidence bands (95%) are shown around the survival curves, and accompanying risk tables provide the number of patients remaining under observation at successive time points.

#### 3.6.2 Early-Onset H/L Patients Not Treated With FOLFOX

In EOCRC H/L patients who did not receive FOLFOX (Figure 2b), JAK-STAT pathway alterations were not associated with a statistically meaningful difference in overall survival (p = 0.25). Although both groups began with similarly high survival probabilities, patients harboring pathway alterations showed a more noticeable decline beginning around 20-30 months, while those without alterations maintained a more gradual downward trajectory. Confidence intervals widened considerably for the altered group, reflecting the limited number of mutation-positive cases contributing to later time points. Despite this visual separation, the overlap in confidence bands indicates that the observed differences are not robust, suggesting that JAK-STAT pathway status does not strongly influence survival outcomes in untreated EOCRC H/L patients.

#### 3.6.3 Late-Onset H/L Patients Treated With FOLFOX

In LOCRC H/L patients receiving FOLFOX (Figure 2c), overall survival did not differ significantly between individuals with and without JAK-STAT pathway alterations (p = 0.24). The two curves tracked closely for much of the follow-up period, with only modest early separation before realigning as time progressed. Patients harboring pathway alterations showed relatively stable survival probabilities, supported by wide confidence intervals reflecting the small number of mutation-positive cases. Meanwhile, the non-altered group demonstrated a gradual decline beginning around 30-40 months but maintained broadly overlapping confidence bands with the altered cohort. Taken together, these findings indicate that JAK-STAT pathway mutation status does not appear to exert a pronounced influence on survival outcomes in LOCRC H/L patients treated with FOLFOX.

#### 3.6.4 Late-Onset H/L Patients Not Treated With FOLFOX

For LOCRC H/L patients who did not receive FOLFOX (Figure 2d), JAK-STAT pathway alterations were not associated with a discernible difference in overall survival (p = 0.31). The survival trajectory of the altered group remained largely stable across the follow-up period, with minimal decline and broad confidence intervals reflecting the very small sample size. In contrast, the non-altered group exhibited a more gradual reduction in survival beginning around 25-30 months, yet the confidence bands of the two groups overlapped extensively. The lack of visible separation between curves, combined with the limited number of altered cases, suggests that JAK-STAT mutation status does not meaningfully influence survival outcomes in untreated LOCRC H/L CRC.

#### 3.6.5 Early-Onset NHW Patients Treated With FOLFOX

In EOCRC NHW patients who received FOLFOX (Figure 2e), JAK-STAT pathway alterations were associated with a statistically significant difference in overall survival (p = 0.00083). Individuals with pathway mutations demonstrated notably better survival, with their curve remaining close to 100% for much of the observation period and showing minimal decline even at extended follow-up. In contrast, patients without alterations exhibited a steady reduction in survival probability that began early and continued progressively across the entire timeframe. The separation between curves increased with time, and the non-overlapping portions of the confidence intervals support the robustness of this difference. These findings indicate that, in EOCRC NHW patients treated with FOLFOX, JAK-STAT pathway mutations may be linked to a favorable survival phenotype.

#### 3.6.6 Early-Onset NHW Patients Not Treated With FOLFOX

In EOCRC NHW patients who did not receive FOLFOX (Figure 2f), JAK-STAT pathway status did not significantly influence overall survival (p = 0.072). Although the curves begin with comparable survival probabilities, a modest separation emerges during mid-follow-up, with the altered group maintaining slightly higher survival estimates. However, the wide and substantially overlapping confidence intervals for both groups indicate considerable uncertainty, particularly among patients with pathway alterations, whose small sample size produced broader variability. By later time points, the trajectories reconverge, suggesting no consistent or durable survival impact attributable to JAK-STAT alterations in this untreated EOCRC NHW population.

#### 3.6.7 Late-Onset NHW Patients Treated With FOLFOX

In LOCRC NHW patients receiving FOLFOX therapy (Figure S1a), JAK-STAT pathway alterations did not exhibit a significant association with overall survival (p = 0.36). Survival curves for altered and non-altered groups remained closely aligned across the follow-up period, with only minor fluctuations in separation that did not persist. Both groups displayed a gradual decline in survival probability over time, and the substantial overlap in their 95% confidence intervals suggests similar outcome trajectories regardless of pathway status. Although the altered cohort showed slightly greater variability, likely attributable to a smaller sample size, there was no indication of a consistent survival advantage or disadvantage linked to JAK-STAT alterations in this LOCRC, FOLFOX-treated NHW population.

#### 3.6.8 Late-Onset NHW Patients Not Treated With FOLFOX

In LOCRC NHW patients who did not receive FOLFOX (Figure S1b), JAK-STAT pathway alterations were associated with a statistically significant difference in overall survival (p = 0.017). Individuals harboring pathway alterations exhibited a more favorable survival pattern, with their curve remaining consistently higher than that of the non-altered group throughout most of the follow-up period. The altered cohort showed a slower decline in survival probability, suggesting a potential protective or less aggressive disease phenotype in the absence of chemotherapy exposure. Although the confidence intervals widened at later time points-reflecting reduced sample sizes-they still maintained partial separation from the non-altered group. By contrast, patients without pathway alterations demonstrated a more rapid and continuous drop in survival, reinforcing the observed divergence. These findings indicate that, in untreated LOCRC NHW patients, JAK-STAT pathway alterations may be linked to improved survival outcomes, warranting further investigation into underlying biological mechanisms.

### 3.7 AI-driven exploratory interrogation of clinical-genomic data prior to formal statistical testing

We began by leveraging the AI-HOPE-JAK-STAT platform to execute a directed post hoc scan of the integrated CRC cohorts, enabling rapid extraction of exploratory signals to shape subsequent statistical testing. Using conversational, query-driven prompts, the system highlighted multiple noteworthy trends.

As an initial step, the AI-HOPE-JAK-STAT (2) agent was used to conduct a rapid exploratory analysis of the integrated CRC datasets, allowing us to identify potential survival-relevant patterns before performing conventional statistical modeling. One representative query examined whether RTK/RAS pathway alterations influenced overall survival specifically among EOCRC NHW patients who had not received FOLFOX therapy (Figure S2). The AI system automatically constructed the comparison groups,185 patients with RTK/RAS pathway alterations and 117 without, and generated a preliminary Kaplan-Meier curve to assess divergence between the two survival profiles. The AI-derived output revealed a notable separation between the altered and non-altered groups, with the altered cohort demonstrating poorer survival across the follow-up period. The survival difference reached statistical significance (log-rank p = 0.0288), suggesting that RTK/RAS dysregulation may adversely affect prognosis in this specific clinical subgroup. Pie-chart visualizations provided by the AI agent confirmed appropriate cohort selection, with altered cases comprising 7.4% of the broader dataset and non-altered controls comprising 4.7%.

As a second exploratory step, the AI-HOPE-JAK-STAT agent was used to systematically screen for clinical and genomic attributes that distinguish EOCRC H/L patients from their NHW counterparts (Figure S3). The agent automatically generated case (H/L, n = 125) and control (NHW, n = 677) cohorts and performed an attribute-by-attribute comparison using natural language-driven prompts and automated contingency testing. The AI system identified a broad set of features that differed significantly between the two groups, encompassing demographic, clinical, and molecular categories. Demographic attributes, such as ethnicity designation and race, showed predictable divergence, while clinical parameters including diagnosis descriptors, cancer type, MSI category, event status, and highest recorded stage also demonstrated significant variation. The platform additionally highlighted disparities in mutation profiles across several well-established CRC driver genes, with notable differences for KRAS, and BRAF. Visualization through stacked bar plots and cohort distribution charts further illustrated the contrast in the proportion of KRAS-mutated samples used in the odds ratio context (8.0 in-context cases vs. 117 in-context controls). Despite the imbalanced sample sizes, the automated statistical testing, based on a 2×2 contingency framework, yielded a highly significant p-value (0.093 for the chi-square test) and an odds ratio of 0.0 (95% CI: 0.045-0.193), indicating a marked disparity in KRAS mutation representation at the exploratory stage.

In a third set of exploratory analysis, the AI-HOPE-JAK-STAT agent evaluated whether BRAF mutations might differ between EOCRC H/L and NHW patients who received FOLFOX therapy (Figure S4). The system automatically assembled a case cohort of EOCRC H/L patients (n = 73) and a control cohort of EOCRC NHW patients (n = 375), applying BRAF mutation status as the contextual attribute for odds ratio testing. Visualization of selected and unselected samples demonstrated that only a very small fraction of the H/L cohort fell within the BRAF-mutated category (0.68%), whereas the proportion was notably higher among NHW patients (7.2%). Fisher’s exact test confirmed the significance of this disparity (p = 0.036). The odds ratio was estimated at 0.0 (95% CI: 0.005-1.464), reflecting the near absence of BRAF-mutated tumors in the H/L subgroup relative to the NHW comparator. These AI-generated findings highlight a substantial ancestry-related divergence in BRAF mutation frequency among EOCRC FOLFOX-treated patients. Although exploratory in nature, the signal suggests that BRAF-driven tumor biology may be far less common in H/L individuals within this clinical context. This distinction prompted further downstream evaluation of ancestry-specific molecular patterns, reinforcing the value of AI-enabled prioritization before formal statistical modeling.

An additional set of exploratory queries using the AI-HOPE-JAK-STAT agent focused on ERBB2 mutation patterns among EOCRC H/L patients stratified by FOLFOX treatment (Figure S5). Although ERBB2 is not classically categorized within the JAK-STAT pathway, there is strong biological justification for interrogating this gene: ERBB2 (HER2) exhibits well-established cross-talk with the JAK-STAT axis. ERBB2 can activate JAK1/JAK2, promote phosphorylation of STAT3/STAT5, and signal through HER2/HER3 heterodimers, thereby influencing downstream processes such as cell survival, angiogenesis, immune evasion, and therapy resistance. In HER2-positive breast and gastric cancers, ERBB2-driven STAT3 activation is a key mechanism contributing to tumor aggressiveness and resistance to targeted therapy. Because of this documented interplay, we intentionally evaluated ERBB2 during hypothesis generation to determine whether similar JAK-STAT, linked biology might emerge in EOCRC H/L. In executing this exploratory task, the AI system automatically assembled the comparison groups, 73 FOLFOX-treated patients and 52 untreated patients, and generated a contingency table to assess whether ERBB2 alterations were enriched in either subgroup. The platform also produced visual summaries, including pie-chart distributions confirming accurate cohort extraction and a stacked bar plot depicting the proportional frequencies of ERBB2-altered versus wild-type samples. Consistent with known CRC biology, preliminary AI-generated statistics revealed that ERBB2 alterations were exceedingly rare in both groups. In the treated cohort, only 0.68% of samples harbored ERBB2 alterations, compared with 1.92% in the untreated group. Fisher’s exact test confirmed the absence of a meaningful difference (p = 0.864), and the odds ratio approximated zero with a wide confidence interval (95% CI: 0.012-10.61), reflecting sparse event counts rather than a true biological disparity. As visualized in the stacked bar plot, both cohorts were dominated by ERBB2-wild-type samples with only isolated altered cases. Collectively, these AI-assisted findings indicate that although ERBB2 was a biologically plausible candidate due to its cross-talk with JAK-STAT, the AI-agent correctly recognized, during the second, more targeted query, that ERBB2 does not demonstrate treatment-related or subgroup-associated enrichment in EOCRC H/L. This underscores the value of AI-HOPE-JAK-STAT in rapidly generating biologically informed hypotheses while also accurately determining when no meaningful signal is present upon confirmatory evaluation.

The exploratory outputs shaped which subgroup comparisons were advanced to formal statistical evaluation. AI-HOPE (2) subsequently automated the filtering of clinical, genomic, and treatment variables to construct analysis-ready cohorts, producing verified mutation tables and survival analyses. This approach reduced manual effort, improved consistency, and accelerated the move from hypothesis generation to statistical confirmation.

## 4. Discussion

This work represents one of the first AI-agent-enabled precision medicine analyst of the JAK-STAT signaling axis in CRC systematically stratified by age at diagnosis, ancestry, and FOLFOX exposure, with a dedicated focus on disproportionately affected H/L populations. Using the AI-HOPE and AI-HOPE=JAK-STAT platforms, we harmonized multi-cohort clinical and genomic data and interrogated pathway-level alterations through natural language-driven analytics. This integrative framework allowed us to uncover context-specific patterns of JAK-STAT dysregulation and related survival effects that would be difficult to detect with conventional, manual curated pipelines.

### Summary of key findings

Several consistent themes emerged. First, JAK-STAT pathway alteration was not uniformly distributed across age, ancestry, and treatment strata. Within the H/L cohort, EOCRC showed a striking treatment-dependent pattern: pathway alterations were significantly more frequent in patients who did not receive FOLFOX than in those who did (21.2% vs. 4.1%; p = 0.003). In contrast, LOCRC H/L cases demonstrated low and relatively similar alteration frequencies regardless of treatment status. Among NHW patients, EOCRC cases displayed comparable alteration rates across treatment groups, but LOCRC NHW tumors harbored significantly more JAK-STAT alteration in the non-FOLFOX group that in those exposed to FOLFOX (13.3% vs. 7.5%; p = 0.0002).

Second, ancestry-stratified comparisons highlighted that early-onset, chemo-naive H/L patients had a substantial higher prevalence of JAK-STAT alterations than their NHW counterparts (21.2% vs. 9.9%; p = 0.002), whereas cross-ancestry differences were attenuated or abstentions in FOLFOX-treated or LOCRC disease. Third, survival analyses revealed that JAK-STAT pathway alterations were generally not prognostic in H/L patients, regardless of age or treatment, but were associated with improved overall survival in several NHW subgroups. NHW EOCRC patients treated with FOLFOX who harbored JAK-STAT mutations experienced significantly better survival than those without alterations (p = 0.0008), and similar protective association was observed in LOCRC NHW patients not treated with FOLFOX (P = 0.017). These results suggest that JAK-STAT alterations may act as ancestry- and treatment-dependent biomarkers, conferring a favorable prognosis in specific NHW context while playing a more neutral role in H/L disease.

### Biological implications of JAK-STAT pathway alterations

The JAK-STAT axis is a central conduit linking inflammatory signals to transcriptional programs that regulate proliferation, survival and immune evasion in CRC Hyperactivation of STAT3 and upstream JAK kinases has been associated with more aggressive phenotypes, chemoresistance, and immunosuppressive microenvironment. However, the net functional impact of individual mutations within this pathway is highly context-dependent, some variants may attenuate signaling, while other enhance oncogenic activity.

In our cohorts, pathway alterations were dominated by missense mutations in genes such as JAK1, JAK3, STAT3, STAT5A, STAT5B, with relatively few truncation or splice-site events. The association between JAK-STAT mutations and improved survival in several NHW subgroups raises the possibility that, in these settings, many alterations may be partial loss-of-function or otherwise deleterious to pro-tumor signaling. Alternatively, JAK-STAT mutations could serve as surrogates for broader immunogenicity or hyper mutated phenotypes that respond more favorably to standard therapies, including FOLFOX. The enrichment of STAT5B mutations specially in untreated NHW EOCRC, further underscores the heterogeneity of the pathway perturbations and suggest gene- and ancestry-specific biology that merits functional follow-up.

### Ancestry-specific genomic patterns and treatment context

Our data highlight important ancestry-linked differences in how JAK-STAT dysregulation manifest across clinical context. The elevated prevalence of pathway alterations in chemo-naive EOCRC H/L tumors relative to NHW EOCRC suggest that inflammation-driven or cytokine-mediated signaling may play a particularly prominent role in early tumorigenesis among H/L individuals. Yet this enrichment did not translate into clear survival penalties or benefits in H/L patients, irrespective of FOLFOX exposure. One explanation is that, in H/L EOCRC, JAK-STAT mutations may occur alongside other aggressive molecular features-such as co-alterations in WNT, RTK/RAS, or PI3K pathways, that dominate prognosis and dilute the independent impact of JAK-STAT status.

In LOCRC NHW, the reduced frequency of JAK-STAT alterations in FOLFOX-treated versus untreated patients could reflect treatment selection biases, differential clinical presentation, or chemotherapy-driven clinal dynamics. For instance, tumors with intact JAK-STAT signaling might be more likely to be selected for intensive therapy based on clinical features, while those harboring pathway mutations may preferentially arise in patients who are less likely to receive FOLFOX. Alternatively, JAK-STAT-mutant clones could be less fit under FOLFOX pressure, resulting in lower post-treatment detection. Although our retrospective design cannot disentangle these possibilities, the observed asymmetries highlight the need to consider both ancestry and treatment history when interpreting pathway alterations.

### Implications for FOLFOX response and prognostic stratification

The most clinically provocative signaling in this study is the association between JAK-STAT alteration and better survival in specific NHW subgroups, particularly EOCRC patients receiving FOLFOX and late0onset patients managed without FOLFOX. These findings contrast with preclinical literature that often links JAK-STAT activation to chemoresistance and poor prognosis, suggesting that the alterations captured in large clinical sequencing cohorts may not uniformly reflect pathway hyper activation. Instead, they may index a subset of tumors with distinct biology, potentially characterized by altered cytokine signaling, enhanced immune recognition, or changes in stratal interactions that render them more susceptible to standard chemotherapy.

In contrast, we did not observe a comparable prognostic effect in H/L patients, despite a higher burden of JAK-STAT alterations in untreated EO disease. This divergence raises the possibility that germline ancestry, environmental exposures, and tumor microenvironmental context modulate the consequences of pathway mutations. If validated in prospective series, JAK-STAT alteration status could be incorporated into ancestry-aware risk models, supporting more nuanced prognostic counseling in NHW patients and motivating additional biomarker discovery efforts in H/L populations, where the pathway’s role appears more complex.

### AI-HOPE-JAK-STAT as an enabling technology

The AI-HOPE and AI-HOPE-JAK-STAT platforms were central to uncovering these patterns. By unifying genomics clinical, and ancestry, and treatment variables into a conversational analytic environment, the system enabled rapid, flexible hypothesis generation ahead of formal statistical modeling. AI-guided queries identified several signals of interest, including the survival disadvantage associated with RTK/RAS alterations in EOCRC NHW patients not treated with FOLFOX. The AI-Agent also surfaced multiple clinical and genomic attributes that distinguished EOCRC H/L form EOCRC NHW patients, such as differences in stage at presentation and key driver mutations.

Crucially, all AI-generated subgroup definitions, frequency patterns, and preliminary statistics were subjected to manual verification and conventional inferential testing. In several instances, AI-flagged hypotheses were not confirmed, underscoring the importance of pairing AI-driven exploration with rigorous biostatistical validation. At the same time, the AI-enabled workflow substantially reduced manual data handling, minimized transcription error, and accelerated the progression from exploratory queries to curated, analysis-ready cohorts, illustrating how conversational agents can function as catalysts rather than replacements for traditional analytics.

### Limitations and future directions

This study has several limitations. Despite the large aggregate cohort, achy stratified subgroups, particularly H/L patients with JAK-STAT alterations, contained relatively few cases, leading to wide confidence intervals and limited power to detect modest effects. Our analysis focused primarily on coding single-nucleotide variants and small indels; we did not systematically incorporate copy-number changes, structural variants, or transcriptional and proteomic readouts that may more directly index pathway activation. Treatment annotations, while sufficient to classify FOLFOX exposure, lacked granular detail on dosing, treatment duration, and use of additional systems agents, which could further modulate outcomes.

The retrospective, multi-cohort design also introduces potential confounding from unmeasured variables such as comorbidities, socioeconomic factors, and center-specific treatment practices, factors that are particularly relevant when interpreting ancestry-related differences. Moreover, the functional impact of individual mutations remains largely inferred rather than experimentally validated; future studies integrating in vitro and in vivo models, as well as single-cell and spatial profiling, will be essential to dissect how specific JAK-STAT alterations reshape tumor-immune interactions across ancestries and treatment contexts.

From a methodological perspective e, AI-HOPE-JAK-STAT depends on the quality of underlying clinical and genomic annotations, systematic biases or misclassifications in source datasets can propagate through AI-assisted analyses. Future iterations of the platform will benefit from explicit integration of data provenance tracking, uncertainty quantification, and fairness auditing to ensure that AI-driven insights do not inadvertently exacerbate existing inequities.

## 5. Conclusion

In summary, this AI-enabled precision oncology study reveals that JAK-STAT pathway alterations in CRC are shaped by a complex interplay of ancestry, age at onset, and FOLFOX exposure. EOCRC H/L patients exhibited a higher burden of JAK-STAT mutations in the absence of FOLFOX, whereas specific NHW subgroups, particularly FOLFOX-treated EOCRC and untreated LOCRC, demonstrated improved survival in the presence of pathway alterations. These findings position JAK-STAT status as a potential ancestry- and treatment-dependent prognostic marker and highlight important biological heterogeneity between H/L and NHW populations.

Beyond these biological insights, our work showcases how conversational AI systems such as AI-HOPE-JAK-STAT can harmonize complex clinical-genomic datasets, accelerate hypothesis generation and guide targeted statistical analyses. By centering disproportionately affected populations and explicitly incorporating ancestry, treatment, and age into pathway-level interrogation, this study illustrates a practical path toward more equitable precision medicine in CRC. Continued integration of AI-driven analytics with mechanistic experimentation and prospect e, ancestrally diverse cohorts will be critical to translating these insights into personalized treatment strategies and improved outcomes for patients historically underrepresented in cancer genomics research.

## Data Availability

All data used in the present study is publicly available at https://www.cbioportal.org/ and https://genie.cbioportal.org. The datasets used in our study were aggregated/summary data, and no individual-level data were used. Additional data can be provided upon reasonable request to the authors.

## Supplementary Materials

**Table S1.** Comparison of Early-Onset Hispanic/Latino (H/L) Patients Treated with FOLFOX versus Not Treated with FOLFOX.

| JAK/STAT Pathway |  |  |  |
| --- | --- | --- | --- |
| Gene | Early-Onset Hispanic/Latino<br>Treated with FOLFOX<br>n (%) | Early-Onset Hispanic/Latino<br>Not Treated with FOLFOX<br>n (%) | p-value |
| JAK1 Mutation |  |  |  |
| Present | 1 (1.4%) | 4 (7.7%) | 0.1594 |
| Absent | 72 (98.6%) | 48 (92.3%) |  |
| JAK2 Mutation |  |  |  |
| Present | 0 (0.0%) | 0 (0.0%) | 1 |
| Absent | 73 (100.0%) | 52 (100.0%) |  |
| JAK3 Mutation |  |  |  |
| Present | 1 (1.4%) | 3 (5.8%) | 0.3067 |
| Absent | 72 (98.6%) | 49 (94.2%) |  |
| SOCS1 Mutation |  |  |  |
| Present | 1 (1.4%) | 0 (0.0%) | 1 |
| Absent | 72 (98.6%) | 52 (100.0%) |  |
| STAT3 Mutation |  |  |  |
| Present | 0 (0.0%) | 2 (3.8%) | 0.1711 |
| Absent | 73 (100.0%) | 50 (96.2%) |  |
| STAT5A Mutation |  |  |  |
| Present | 0 (0.0%) | 2 (3.8%) | 0.1711 |
| Absent | 73 (100.0%) | 50 (96.2%) |  |
| STAT5B Mutation |  |  |  |
| Present | 0 (0.0%) | 5 (9.6%) | 0.01108 |
| Absent | 73 (100.0%) | 47 (90.4%) |  |

**Table S2.** Comparison of Late-Onset Hispanic/Latino (H/L) Patients Treated with FOLFOX versus Not Treated with FOLFOX.

| JAK/STAT Pathway |  |  |  |
| --- | --- | --- | --- |
| Gene | Late-Onset Hispanic/Latino<br>Treated with FOLFOX<br>n (%) | Late-Onset Hispanic/Latino<br>Not Treated with FOLFOX<br>n (%) | p-value |
| JAK1 Mutation |  |  |  |
| Present | 2 (2.2%) | 2 (2.2%) | 1 |
| Absent | 89 (97.8%) | 89 (97.8%) |  |
| JAK2 Mutation |  |  |  |
| Present | 2 (2.2%) | 2 (2.2%) | 1 |
| Absent | 89 (97.8%) | 89 (97.8%) |  |
| JAK3 Mutation |  |  |  |
| Present | 4 (4.4%) | 4 (4.4%) | 1 |
| Absent | 87 (95.6%) | 87 (95.6%) |  |
| SOCS1 Mutation |  |  |  |
| Present | 0 (0.0%) | 0 (0.0%) | 1 |
| Absent | 91 (100.0%) | 91 (100.0%) |  |
| STAT3 Mutation |  |  |  |
| Present | 1 (1.1%) | 1 (1.1%) | 1 |
| Absent | 90 (98.9%) | 90 (98.9%) |  |
| STAT5A Mutation |  |  |  |
| Present | 2 (2.2%) | 2 (2.2%) | 1 |
| Absent | 89 (97.8%) | 89 (97.8%) |  |
| STAT5B Mutation |  |  |  |
| Present | 1 (1.1%) | 1 (1.1%) | 1 |
| Absent | 90 (98.9%) | 90 (98.9%) |  |

**Table S3.** Comparison of Early-Onset Non-Hispanic White (NHW) Patients Treated with FOLFOX versus Not Treated with FOLFOX.

| JAK/STAT Pathway |  |  |  |
| --- | --- | --- | --- |
| Gene | Early-Onset NHW<br>Treated with FOLFOX<br>n (%) | Early-Onset NHW<br>Not Treated with FOLFOX<br>n (%) | p-value |
| JAK1 Mutation |  |  |  |
| Present | 10 (2.7%) | 10 (3.3%) | 0.7917 |
| Absent | 365 (97.3%) | 292 (96.7%) |  |
| JAK2 Mutation |  |  |  |
| Present | 5 (1.3%) | 9 (3.0%) | 0.2206 |
| Absent | 370 (98.7%) | 293 (97.0%) |  |
| JAK3 Mutation |  |  |  |
| Present | 4 (1.1%) | 14 (4.6%) | 0.006502 |
| Absent | 371 (98.9%) | 288 (95.4%) |  |
| SOCS1 Mutation |  |  |  |
| Present | 1 (0.3%) | 1 (0.3%) | 1 |
| Absent | 374 (99.7%) | 301 (99.7%) |  |
| STAT3 Mutation |  |  |  |
| Present | 7 (1.9%) | 3 (1.0%) | 0.5245 |
| Absent | 368 (98.1%) | 299 (99.0%) |  |
| STAT5A Mutation |  |  |  |
| Present | 3 (0.8%) | 5 (1.7%) | 0.4772 |
| Absent | 372 (99.2%) | 297 (98.3%) |  |
| STAT5B Mutation |  |  |  |
| Present | 5 (1.3%) | 7 (2.3%) | 0.5015 |
| Absent | 370 (98.7%) | 295 (97.7%) |  |

**Table S4.** Comparison of Late-Onset Non-Hispanic White (NHW) Patients Treated with FOLFOX versus Not Treated with FOLFOX.

| JAK/STAT Pathway |  |  |  |
| --- | --- | --- | --- |
| Gene | Late-Onset NHW<br>Treated with FOLFOX<br>n (%) | Late-Onset NHW<br>Not Treated with FOLFOX<br>n (%) | p-value |
| JAK1 Mutation |  |  |  |
| Present | 10 (1.8%) | 39 (6.0%) | 0.1151 |
| Absent | 292 (98.2%) | 614 (94.0%) |  |
| JAK2 Mutation |  |  |  |
| Present | 9 (1.2%) | 17 (2.6%) | 0.9054 |
| Absent | 293 (98.8%) | 636 (97.4%) |  |
| JAK3 Mutation |  |  |  |
| Present | 14 (2.6%) | 28 (4.3%) | 0.9409 |
| Absent | 288 (97.4%) | 625 (95.7%) |  |
| SOCS1 Mutation |  |  |  |
| Present | 1 (0.4%) | 2 (0.3%) | 1 |
| Absent | 301 (99.6%) | 651 (99.7%) |  |
| STAT3 Mutation |  |  |  |
| Present | 3 (1.5%) | 11 (1.7%) | 0.5666 |
| Absent | 299 (98.5%) | 642 (98.3%) |  |
| STAT5A Mutation |  |  |  |
| Present | 5 (0.7%) | 9 (1.4%) | 0.9664 |
| Absent | 297 (99.3%) | 644 (98.6%) |  |
| STAT5B Mutation |  |  |  |
| Present | 7 (0.9%) | 21 (3.2%) | 0.5764 |
| Absent | 295 (99.1%) | 632 (96.8%) |  |

**Table S5.** Comparison of Early-Onset versus Late-Onset Hispanic/Latino (H/L) Patients Treated with FOLFOX.

| JAK/STAT Pathway |  |  |  |
| --- | --- | --- | --- |
| Gene | Early-Onset Hispanic/Latino<br>Treated with FOLFOX<br>n (%) | Late-Onset Hispanic/Latino<br>Treated with FOLFOX<br>n (%) | p-value |
| JAK1 Mutation |  |  |  |
| Present | 1 (1.4%) | 2 (2.2%) | 1 |
| Absent | 72 (98.6%) | 89 (97.8%) |  |
| JAK2 Mutation |  |  |  |
| Present | 0 (0.0%) | 2 (2.2%) | 0.503 |
| Absent | 73 (100.0%) | 89 (97.8%) |  |
| JAK3 Mutation |  |  |  |
| Present | 1 (1.4%) | 4 (4.4%) | 0.3826 |
| Absent | 72 (98.6%) | 87 (95.6%) |  |
| SOCS1 Mutation |  |  |  |
| Present | 1 (1.4%) | 0 (0.0%) | 0.4451 |
| Absent | 72 (98.6%) | 91 (100.0%) |  |
| STAT3 Mutation |  |  |  |
| Present | 0 (0.0%) | 1 (1.1%) | 1 |
| Absent | 73 (100.0%) | 90 (98.9%) |  |
| STAT5A Mutation |  |  |  |
| Present | 0 (0.0%) | 2 (2.2%) | 0.503 |
| Absent | 73 (100.0%) | 89 (97.8%) |  |
| STAT5B Mutation |  |  |  |
| Present | 0 (0.0%) | 1 (1.1%) | 1 |
| Absent | 73 (100.0%) | 90 (98.9%) |  |

**Table S6.** Comparison of Early-Onset versus Late-Onset Hispanic/Latino (H/L) Patients Not Treated with FOLFOX.

| JAK/STAT Pathway |  |  |  |
| --- | --- | --- | --- |
| Gene | Early-Onset Hispanic/Latino<br>Not Treated with FOLFOX<br>n (%) | Late-Onset Hispanic/Latino<br>Not Treated with FOLFOX<br>n (%) | p-value |
| JAK1 Mutation |  |  |  |
| Present | 4 (7.7%) | 2 (2.2%) | 0.1903 |
| Absent | 48 (92.3%) | 89 (97.8%) |  |
| JAK2 Mutation |  |  |  |
| Present | 0 (0.0%) | 2 (2.2%) | 0.5339 |
| Absent | 52 (100.0%) | 89 (97.8%) |  |
| JAK3 Mutation |  |  |  |
| Present | 3 (5.8%) | 4 (4.4%) | 0.705 |
| Absent | 49 (94.2%) | 87 (95.6%) |  |
| SOCS1 Mutation |  |  |  |
| Present | 0 (0.0%) | 0 (0.0%) | 1 |
| Absent | 52 (100.0%) | 91 (100.0%) |  |
| STAT3 Mutation |  |  |  |
| Present | 2 (3.8%) | 1 (1.1%) | 0.2992 |
| Absent | 50 (96.2%) | 90 (98.9%) |  |
| STAT5A Mutation |  |  |  |
| Present | 2 (3.8%) | 2 (2.2%) | 0.6218 |
| Absent | 50 (96.2%) | 89 (97.8%) |  |
| STAT5B Mutation |  |  |  |
| Present | 5 (9.6%) | 1 (1.1%) | 0.02405 |
| Absent | 47 (90.4%) | 90 (98.9%) |  |

**Table S7.**
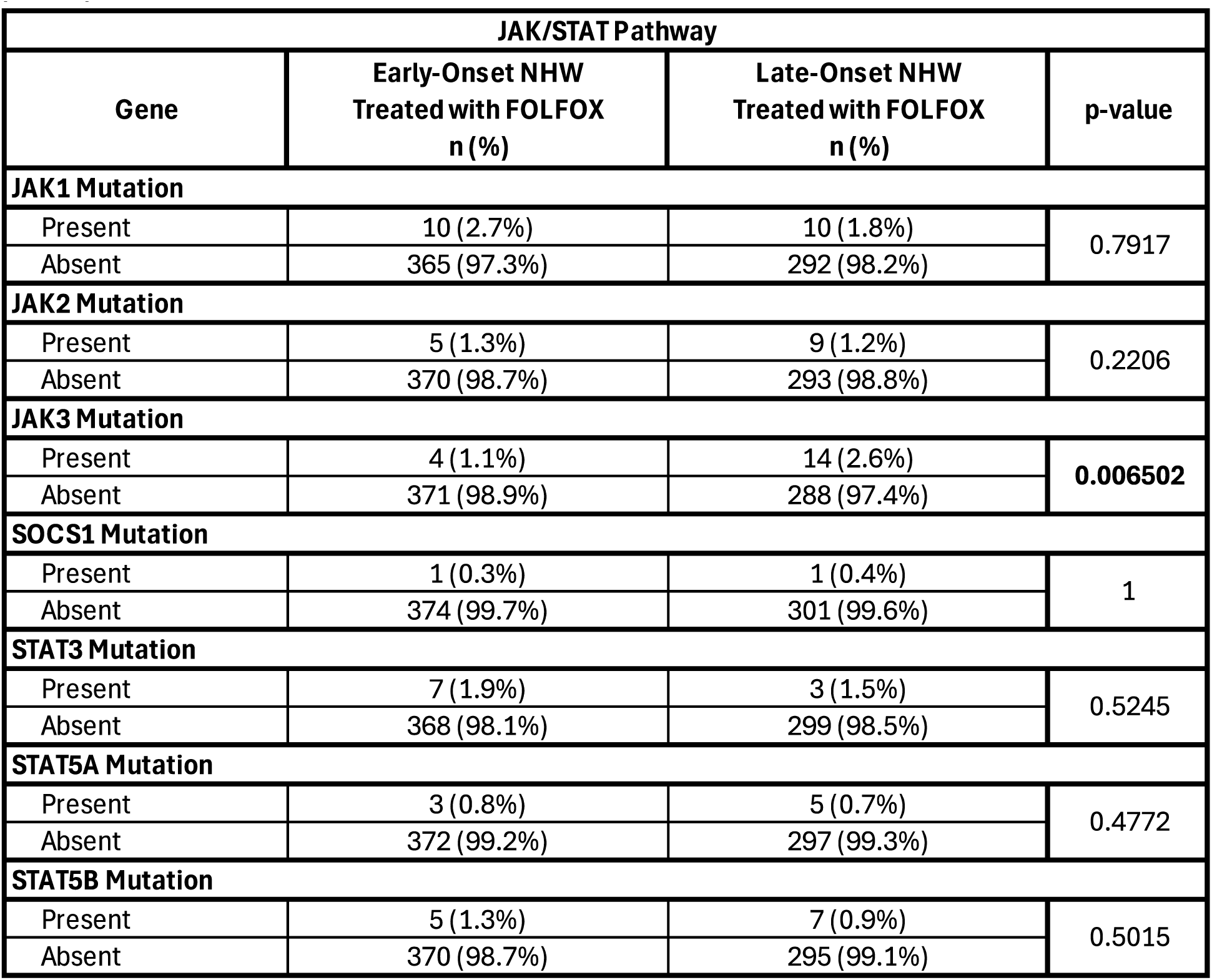
Comparison of Early-Onset versus Late-Onset Non-Hispanic White (NHW) Patients Treated with FOLFOX.

**Table S8.** Comparison of Early-Onset Hispanic/Latino (H/L) versus Early-Onset Non-Hispanic White (NHW) Patients Treated with FOLFOX.

| JAK/STAT Pathway |  |  |  |
| --- | --- | --- | --- |
| Gene | Early-Onset Hispanic/Latino<br>Treated with FOLFOX<br>n (%) | Early-Onset NHW<br>Treated with FOLFOX<br>n (%) | p-value |
| JAK1 Mutation |  |  |  |
| Present | 1 (1.4%) | 10 (2.7%) | 1 |
| Absent | 72 (98.6%) | 365 (97.3%) |  |
| JAK2 Mutation |  |  |  |
| Present | 0 (0.0%) | 5 (1.3%) | 1 |
| Absent | 73 (100.0%) | 370 (98.7%) |  |
| JAK3 Mutation |  |  |  |
| Present | 1 (1.4%) | 4 (1.1%) | 0.5909 |
| Absent | 72 (98.6%) | 371 (98.9%) |  |
| SOCS1 Mutation |  |  |  |
| Present | 1 (1.4%) | 1 (0.3%) | 0.2996 |
| Absent | 72 (98.6%) | 374 (99.7%) |  |
| STAT3 Mutation |  |  |  |
| Present | 0 (0.0%) | 7 (1.9%) | 0.6049 |
| Absent | 73 (100.0%) | 368 (98.1%) |  |
| STAT5A Mutation |  |  |  |
| Present | 0 (0.0%) | 3 (0.8%) | 1 |
| Absent | 73 (100.0%) | 372 (99.2%) |  |
| STAT5B Mutation |  |  |  |
| Present | 0 (0.0%) | 5 (1.3%) | 1 |
| Absent | 73 (100.0%) | 370 (98.7%) |  |

**Table S9.** Comparison of Early-Onset Hispanic/Latino (H/L) versus Early-Onset Non-Hispanic White (NHW) Patients Not Treated with FOLFOX.

| JAK/STAT Pathway |  |  |  |
| --- | --- | --- | --- |
| Gene | Early-Onset Hispanic/Latino<br>Not Treated with FOLFOX<br>n (%) | Early-Onset NHW<br>Not Treated with FOLFOX<br>n (%) | p-value |
| JAK1 Mutation |  |  |  |
| Present | 4 (7.7%) | 10 (3.3%) | 0.1342 |
| Absent | 48 (92.3%) | 292 (96.7%) |  |
| JAK2 Mutation |  |  |  |
| Present | 0 (0.0%) | 9 (3.0%) | 0.3669 |
| Absent | 52 (100.0%) | 293 (97.0%) |  |
| JAK3 Mutation |  |  |  |
| Present | 3 (5.8%) | 14 (4.6%) | 0.7245 |
| Absent | 49 (94.2%) | 288 (95.4%) |  |
| SOCS1 Mutation |  |  |  |
| Present | 0 (0.0%) | 1 (0.3%) | 1 |
| Absent | 52 (100.0%) | 301 (99.7%) |  |
| STAT3 Mutation |  |  |  |
| Present | 2 (3.8%) | 3 (1.0%) | 0.158 |
| Absent | 50 (96.2%) | 299 (99.0%) |  |
| STAT5A Mutation |  |  |  |
| Present | 2 (3.8%) | 5 (1.7%) | 0.2743 |
| Absent | 50 (96.2%) | 297 (98.3%) |  |
| STAT5B Mutation |  |  |  |
| Present | 5 (9.6%) | 7 (2.3%) | 0.01994 |
| Absent | 47 (90.4%) | 295 (97.7%) |  |

**Table S10.**
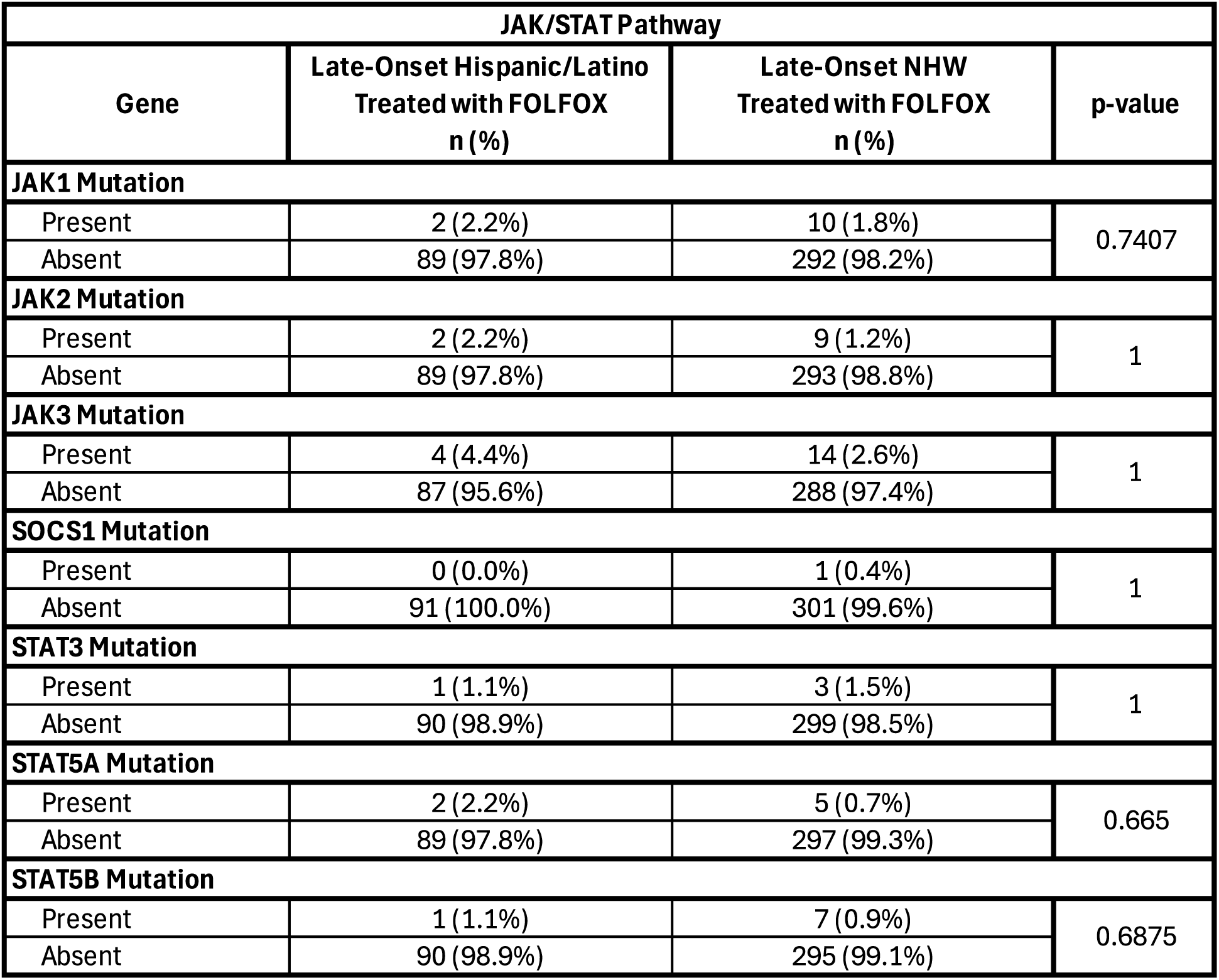
Comparison of Late-Onset Hispanic/Latino (H/L) versus Late-Onset Non-Hispanic White (NHW) Patients Treated with FOLFOX.

**Table S11.**
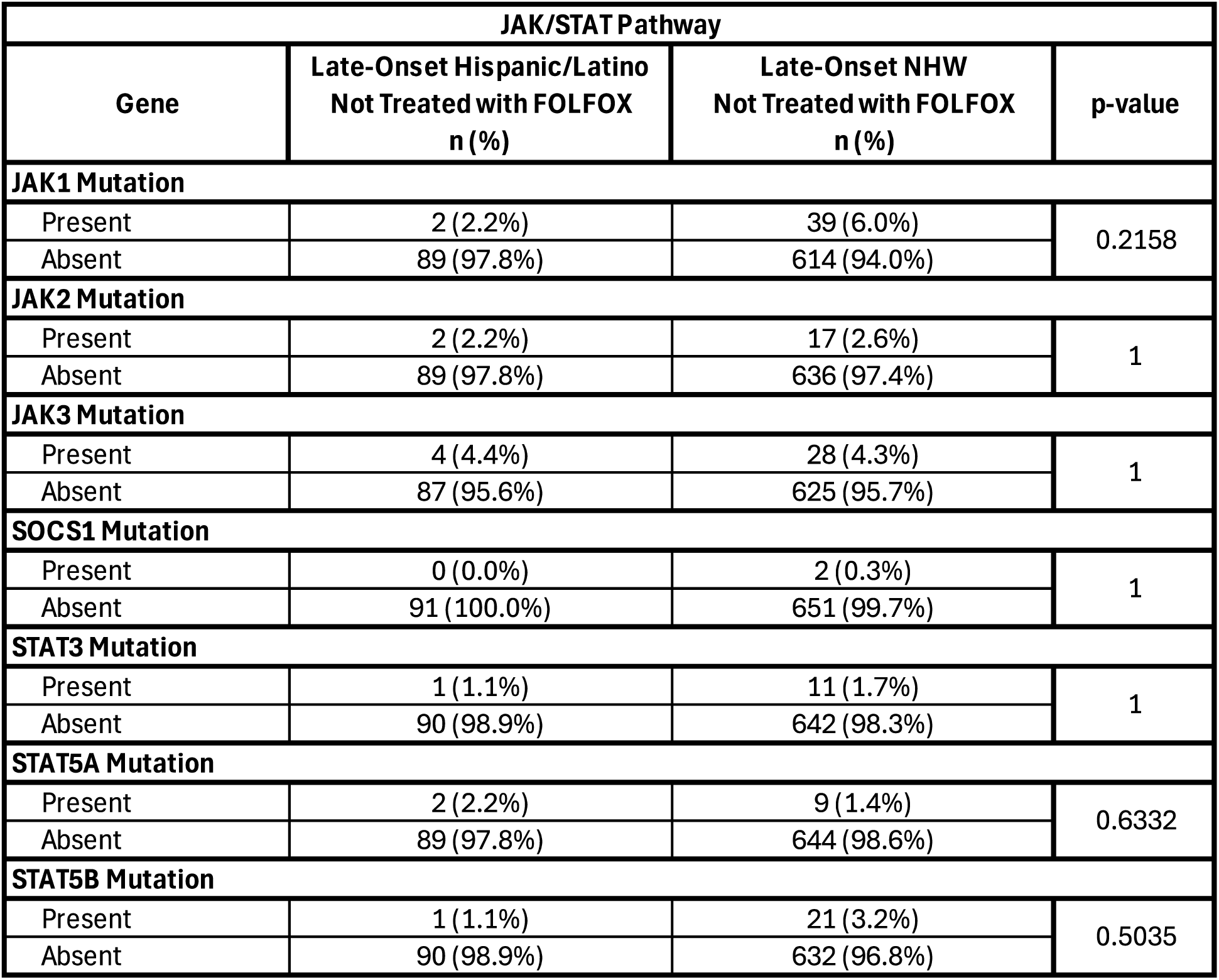
Comparison of Late-Onset Hispanic/Latino (H/L) versus Late-Onset Non-Hispanic White (NHW) Patients Not Treated with FOLFOX.

| JAK/STAT Pathway |  |  |  |
| --- | --- | --- | --- |
| Gene | Late-Onset Hispanic/Latino<br>Not Treated with FOLFOX<br>n (%) | Late-Onset NHW<br>Not Treated with FOLFOX<br>n (%) | p-value |
| JAK1 Mutation |  |  |  |
| Present | 2 (2.2%) | 39 (6.0%) | 0.2158 |
| Absent | 89 (97.8%) | 614 (94.0%) |  |
| JAK2 Mutation |  |  |  |
| Present | 2 (2.2%) | 17 (2.6%) | 1 |
| Absent | 89 (97.8%) | 636 (97.4%) |  |
| JAK3 Mutation |  |  |  |
| Present | 4 (4.4%) | 28 (4.3%) | 1 |
| Absent | 87 (95.6%) | 625 (95.7%) |  |
| SOCS1 Mutation |  |  |  |
| Present | 0 (0.0%) | 2 (0.3%) | 1 |
| Absent | 91 (100.0%) | 651 (99.7%) |  |
| STAT3 Mutation |  |  |  |
| Present | 1 (1.1%) | 11 (1.7%) | 1 |
| Absent | 90 (98.9%) | 642 (98.3%) |  |
| STAT5A Mutation |  |  |  |
| Present | 2 (2.2%) | 9 (1.4%) | 0.6332 |
| Absent | 89 (97.8%) | 644 (98.6%) |  |
| STAT5B Mutation |  |  |  |
| Present | 1 (1.1%) | 21 (3.2%) | 0.5035 |
| Absent | 90 (98.9%) | 632 (96.8%) |  |

**Table S12.**
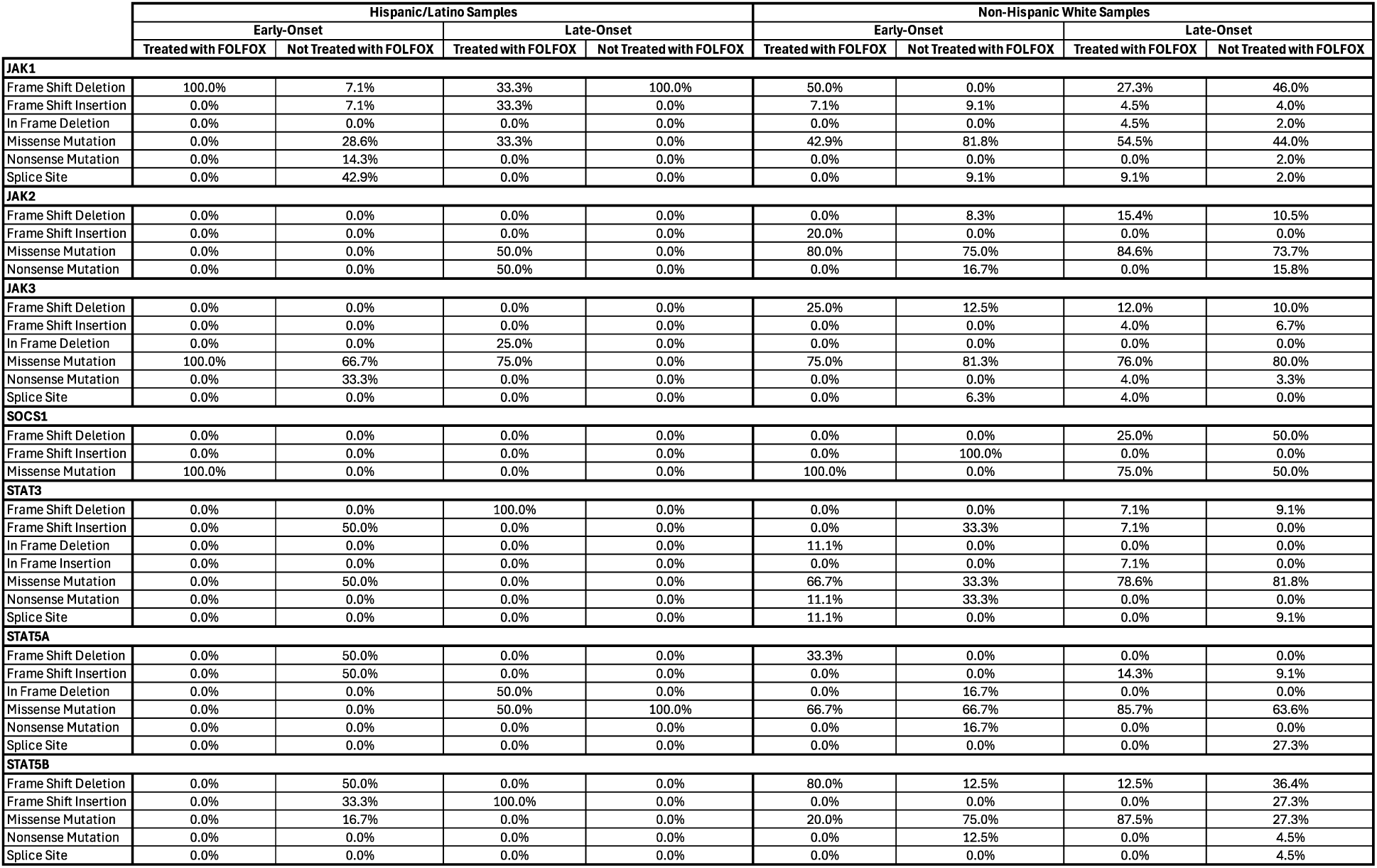
Patterns of JAK-STAT Pathway Mutation Types Across Ancestry, Age-of-Onset Groups, and FOLFOX Treatment in Colorectal Cancer. This table summarizes the distribution of mutation types observed in JAK-STAT pathway genes, categorized by ancestry [Hispanic/Latino (H/L) and Non-Hispanic White (NHW)], diagnostic age group [early-onset (EO) vs. late-onset (LO)], and FOLFOX treatment exposure (treated vs. untreated). Mutation classes include frameshift insertions and deletions, in-frame indels, missense and nonsense mutations, nonstop variants, splice site and splice-region alterations, and changes affecting translation initiation. The percentages shown represent the relative contribution of each mutation class to the total mutations identified for each gene within the specified subgroup. This table provides a comparative view of variation in mutation spectra across demographic and clinical strata, enabling assessment of how ancestry, age, and treatment status relate to differences in the types of JAK-STAT alterations detected.

|  | Hispanic/Latino Samples |  |  |  |  |  | Non-Hispanic White Samples |  |  |  |  |
| --- | --- | --- | --- | --- | --- | --- | --- | --- | --- | --- | --- |
|  | Early-Onset |  |  | Late-Onset |  |  | Early-Onset |  |  | Late-Onset |  |
|  | Treated with FOLFOX | Not Treated with FOLFOX |  | Treated with FOLFOX | Not Treated with FOLFOX |  | Treated with FOLFOX | Not Treated with FOLFOX |  | Treated with FOLFOX | Not Treated with FOLFOX |
| JAK1 |  |  |  |  |  |  |  |  |  |  |  |
| Frame Shift Deletion | 100.0% | 7.1% |  | 33.3% | 100.0% |  | 50.0% | 0.0% |  | 27.3% | 46.0% |
| Frame Shift Insertion | 0.0% | 7.1% |  | 33.3% | 0.0% |  | 7.1% | 9.1% |  | 4.5% | 4.0% |
| In Frame Deletion | 0.0% | 0.0% |  | 0.0% | 0.0% |  | 0.0% | 0.0% |  | 4.5% | 2.0% |
| Missense Mutation | 0.0% | 28.6% |  | 33.3% | 0.0% |  | 42.9% | 81.8% |  | 54.5% | 44.0% |
| Nonsense Mutation | 0.0% | 14.3% |  | 0.0% | 0.0% |  | 0.0% | 0.0% |  | 0.0% | 2.0% |
| Splice Site | 0.0% | 42.9% |  | 0.0% | 0.0% |  | 0.0% | 9.1% |  | 9.1% | 2.0% |
| JAK2 |  |  |  |  |  |  |  |  |  |  |  |
| Frame Shift Deletion | 0.0% | 0.0% |  | 0.0% | 0.0% |  | 0.0% | 8.3% |  | 15.4% | 10.5% |
| Frame Shift Insertion | 0.0% | 0.0% |  | 0.0% | 0.0% |  | 20.0% | 0.0% |  | 0.0% | 0.0% |
| Missense Mutation | 0.0% | 0.0% |  | 50.0% | 0.0% |  | 80.0% | 75.0% |  | 84.6% | 73.7% |
| Nonsense Mutation | 0.0% | 0.0% |  | 50.0% | 0.0% |  | 0.0% | 16.7% |  | 0.0% | 15.8% |
| JAK3 |  |  |  |  |  |  |  |  |  |  |  |
| Frame Shift Deletion | 0.0% | 0.0% |  | 0.0% | 0.0% |  | 25.0% | 12.5% |  | 12.0% | 10.0% |
| Frame Shift Insertion | 0.0% | 0.0% |  | 0.0% | 0.0% |  | 0.0% | 0.0% |  | 4.0% | 6.7% |
| In Frame Deletion | 0.0% | 0.0% |  | 25.0% | 0.0% |  | 0.0% | 0.0% |  | 0.0% | 0.0% |
| Missense Mutation | 100.0% | 66.7% |  | 75.0% | 0.0% |  | 75.0% | 81.3% |  | 76.0% | 80.0% |
| Nonsense Mutation | 0.0% | 33.3% |  | 0.0% | 0.0% |  | 0.0% | 0.0% |  | 4.0% | 3.3% |
| Splice Site | 0.0% | 0.0% |  | 0.0% | 0.0% |  | 0.0% | 6.3% |  | 4.0% | 0.0% |
| SOCS1 |  |  |  |  |  |  |  |  |  |  |  |
| Frame Shift Deletion | 0.0% | 0.0% |  | 0.0% | 0.0% |  | 0.0% | 0.0% |  | 25.0% | 50.0% |
| Frame Shift Insertion | 0.0% | 0.0% |  | 0.0% | 0.0% |  | 0.0% | 100.0% |  | 0.0% | 0.0% |
| Missense Mutation | 100.0% | 0.0% |  | 0.0% | 0.0% |  | 100.0% | 0.0% |  | 75.0% | 50.0% |
| STAT3 |  |  |  |  |  |  |  |  |  |  |  |
| Frame Shift Deletion | 0.0% | 0.0% |  | 100.0% | 0.0% |  | 0.0% | 0.0% |  | 7.1% | 9.1% |
| Frame Shift Insertion | 0.0% | 50.0% |  | 0.0% | 0.0% |  | 0.0% | 33.3% |  | 7.1% | 0.0% |
| In Frame Deletion | 0.0% | 0.0% |  | 0.0% | 0.0% |  | 11.1% | 0.0% |  | 0.0% | 0.0% |
| In Frame Insertion | 0.0% | 0.0% |  | 0.0% | 0.0% |  | 0.0% | 0.0% |  | 7.1% | 0.0% |
| Missense Mutation | 0.0% | 50.0% |  | 0.0% | 0.0% |  | 66.7% | 33.3% |  | 78.6% | 81.8% |
| Nonsense Mutation | 0.0% | 0.0% |  | 0.0% | 0.0% |  | 11.1% | 33.3% |  | 0.0% | 0.0% |
| Splice Site | 0.0% | 0.0% |  | 0.0% | 0.0% |  | 11.1% | 0.0% |  | 0.0% | 9.1% |
| STAT5A |  |  |  |  |  |  |  |  |  |  |  |
| Frame Shift Deletion | 0.0% | 50.0% |  | 0.0% | 0.0% |  | 33.3% | 0.0% |  | 0.0% | 0.0% |
| Frame Shift Insertion | 0.0% | 50.0% |  | 0.0% | 0.0% |  | 0.0% | 0.0% |  | 14.3% | 9.1% |
| In Frame Deletion | 0.0% | 0.0% |  | 50.0% | 0.0% |  | 0.0% | 16.7% |  | 0.0% | 0.0% |
| Missense Mutation | 0.0% | 0.0% |  | 50.0% | 100.0% |  | 66.7% | 66.7% |  | 85.7% | 63.6% |
| Nonsense Mutation | 0.0% | 0.0% |  | 0.0% | 0.0% |  | 0.0% | 16.7% |  | 0.0% | 0.0% |
| Splice Site | 0.0% | 0.0% |  | 0.0% | 0.0% |  | 0.0% | 0.0% |  | 0.0% | 27.3% |
| STAT5B |  |  |  |  |  |  |  |  |  |  |  |
| Frame Shift Deletion | 0.0% | 50.0% |  | 0.0% | 0.0% |  | 80.0% | 12.5% |  | 12.5% | 36.4% |
| Frame Shift Insertion | 0.0% | 33.3% |  | 100.0% | 0.0% |  | 0.0% | 0.0% |  | 0.0% | 27.3% |
| Missense Mutation | 0.0% | 16.7% |  | 0.0% | 0.0% |  | 20.0% | 75.0% |  | 87.5% | 27.3% |
| Nonsense Mutation | 0.0% | 0.0% |  | 0.0% | 0.0% |  | 0.0% | 12.5% |  | 0.0% | 4.5% |
| Splice Site | 0.0% | 0.0% |  | 0.0% | 0.0% |  | 0.0% | 0.0% |  | 0.0% | 4.5% |

**Figure S1.**
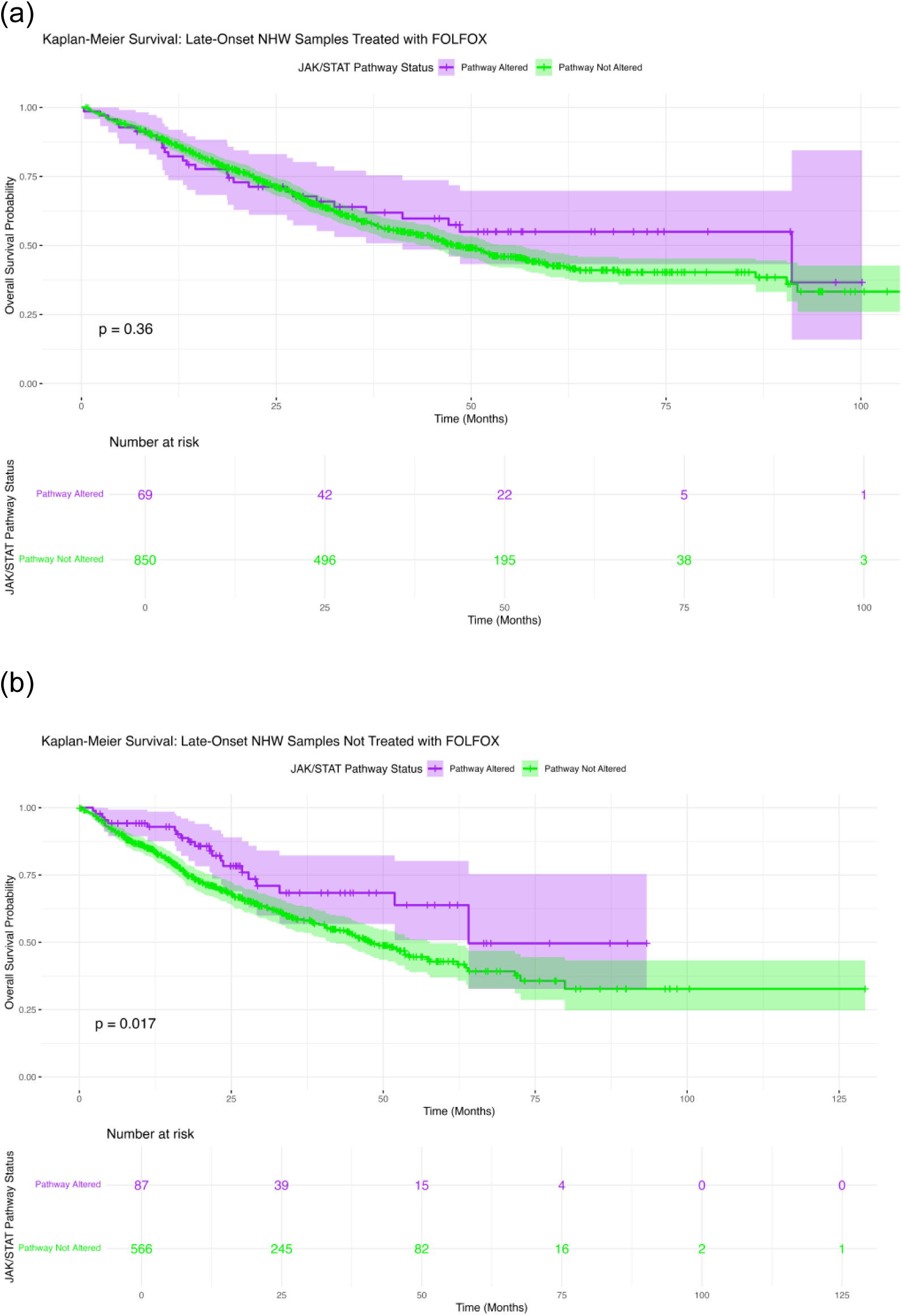
Kaplan-Meier survival curves evaluating the impact of JAK-STAT pathway alterations on overall survival in colorectal cancer (CRC), stratified by age group, ancestry, and FOLFOX treatment. Survival outcomes are presented for two subgroups: (a) late-onset NHW patients who received FOLFOX therapy, and (b) late-onset NHW patients who did not receive FOLFOX.

**Figure S2.**
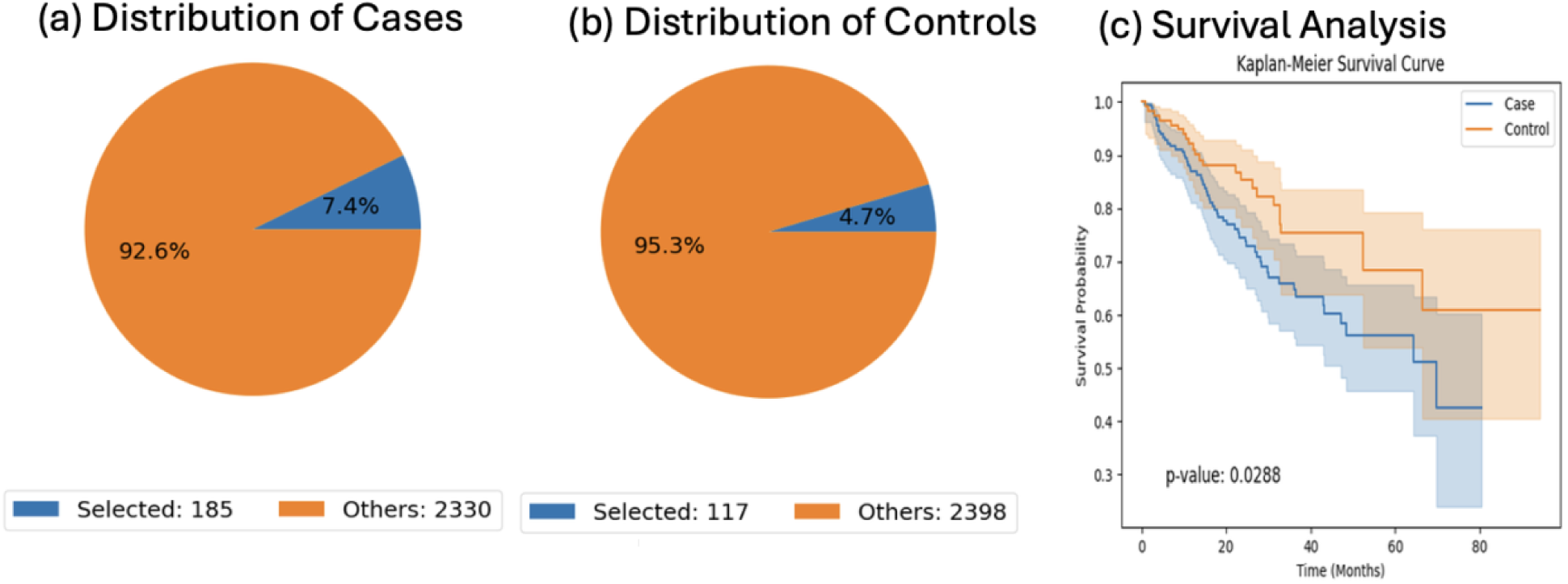
AI-guided cohort construction and survival comparison for early-onset Non-Hispanic White (NHW) colorectal cancer (CRC) patients not treated with FOLFOX, stratified by RTK/RAS pathway alteration status. Using natural language-driven querying, the AI-HOPE-JAK-STAT platform automatically identified case and control cohorts based on user-defined clinical and genomic criteria. (a) The case cohort consisted of early-onset (EO) NHW CRC patients not treated with FOLFOX who harbored RTK/RAS pathway alterations (n = 185), representing 7.4% of all samples within this subgroup. (b) The control cohort included EO NHW patients not treated with FOLFOX and lacking RTK/RAS pathway alterations (n = 117), corresponding to 4.7% of samples in this category. (c) Kaplan-Meier overall survival (OS) analysis comparing altered versus non-altered groups revealed a statistically significant difference (log-rank p = 0.0288), with pathway-altered patients demonstrating reduced survival probabilities over time. Shaded regions indicate 95% confidence intervals. These AI-derived cohort selections and survival trends highlight the potential prognostic relevance of RTK/RAS pathway alterations in EO NHW patients who did not receive FOLFOX.

**Figure S3.**
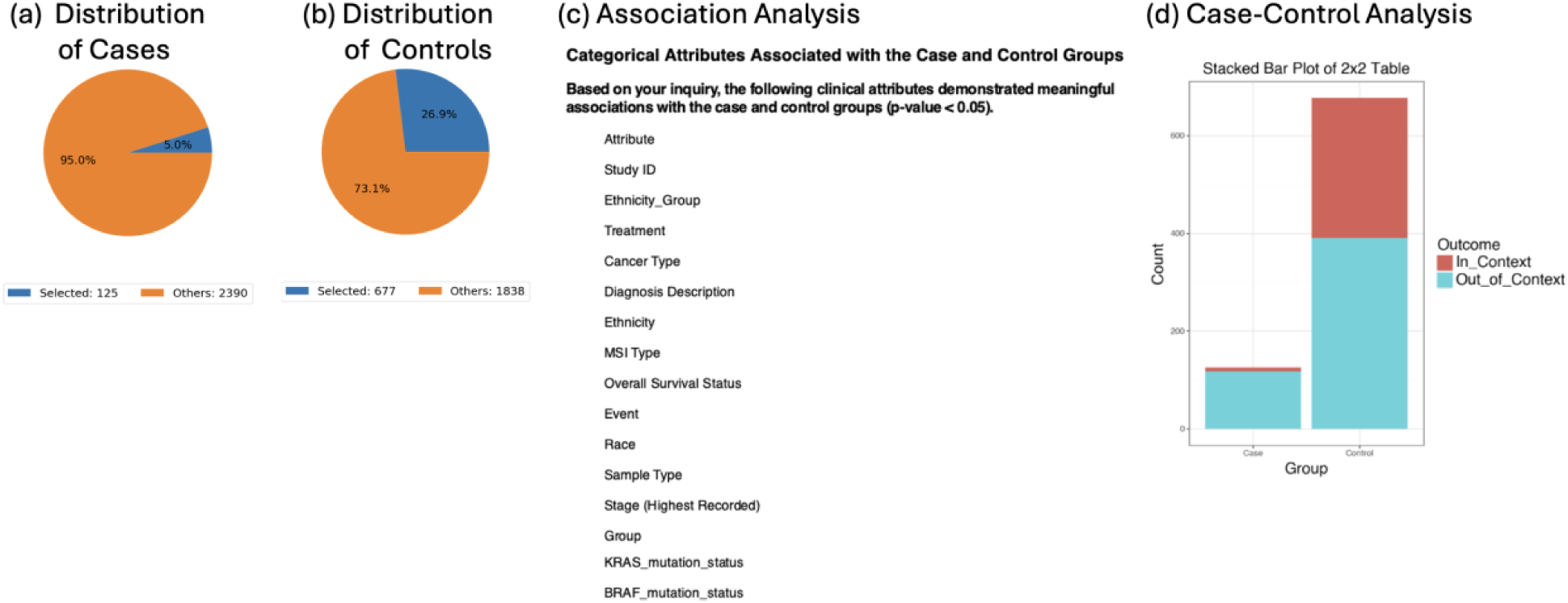
AI-guided identification of clinical attributes distinguishing case and control cohorts in the AI-HOPE-JAK-STAT exploratory analysis. Using the AI-HOPE-JAK-STAT platform, case samples (n = 125) were compared with control samples (n = 677) to identify categorical clinical features meaningfully associated with cohort membership. Panels (a) and (b) display the distribution of selected (in-context) versus unselected samples within the case and control groups, respectively. The case cohort contained 95.0% unselected samples and 5.0% selected samples, whereas the control cohort contained 73.1% unselected and 26.9% selected samples, illustrating a higher enrichment of selected samples within the control group. Panel (c) summarizes the categorical attributes identified by the AI-HOPE-JAK-STAT system as significantly associated with case-control classification (p < 0.05). These attributes included Study ID, Ethnicity_Group, Treatment, Cancer Type, Diagnosis Description, MSI Type, Overall Survival Status, Event, Race, Sample Type, Stage (highest recorded), Group, and key mutation variables such as KRAS and BRAF mutation status. These associations provide data-driven hypotheses regarding clinical and molecular variables contributing to differences between the two groups. Panel (d) shows the stacked 2×2 bar plot comparing the distribution of in-context and out-of-context samples across the case and control cohorts. This visualization highlights the disproportionate representation of selected samples within the control group, suggesting potentially meaningful clinical or molecular distinctions requiring further confirmatory statistical evaluation. Together, these AI-guided exploratory findings demonstrate the capacity of AI-HOPE-JAK-STAT to rapidly surface clinically relevant categorical differences between cohorts, supporting downstream hypothesis generation and deeper statistical modeling.

**Figure S4.**
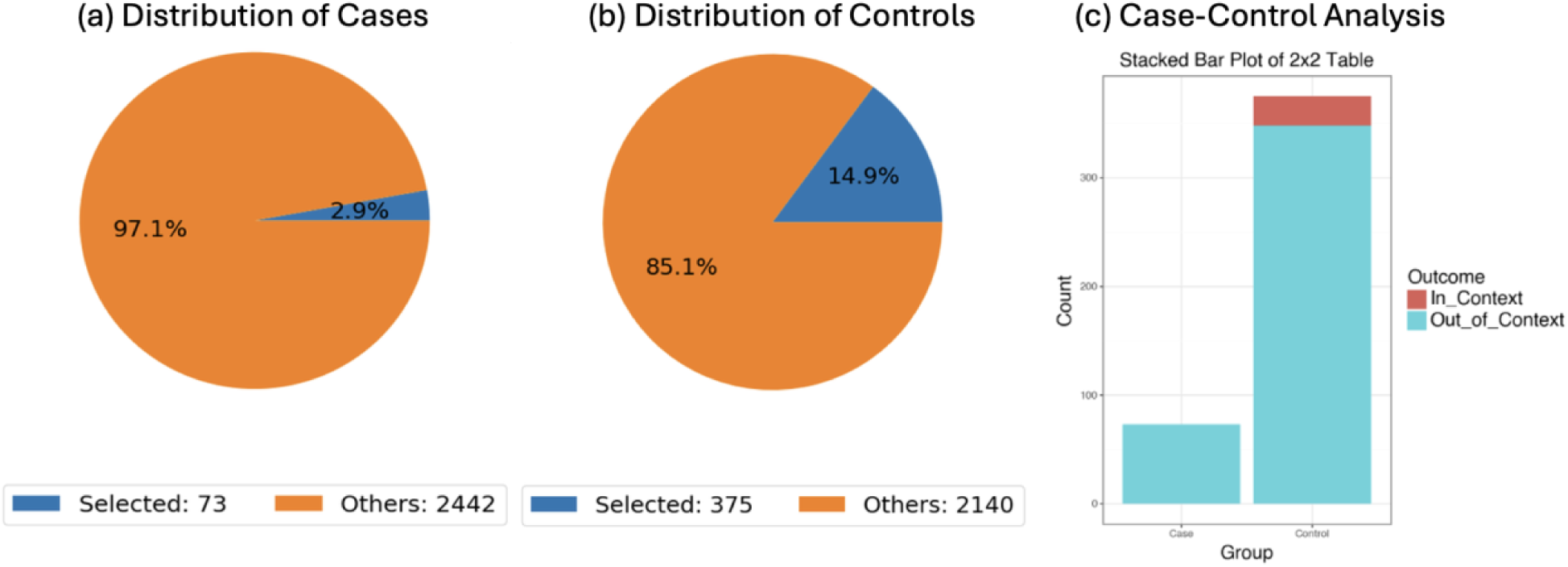
AI-assisted comparison of BRAF mutation prevalence between early-onset Hispanic/Latino (H/L) and Non-Hispanic White (NHW) colorectal cancer (CRC) patients treated with FOLFOX. Using the AI-HOPE-JAK-STAT platform, early-onset H/L CRC patients (case cohort; n = 73) were compared with early-onset NHW patients (control cohort; n = 375) to evaluate differences in BRAF mutation frequency under identical treatment conditions. The pie charts illustrate the proportion of selected samples (BRAF-mutated; blue) versus unselected samples (non-mutated; orange) in each cohort, revealing a substantially lower fraction of BRAF-mutant cases in the H/L group. The stacked bar plot on the right visualizes “In_Context” (BRAF-mutated) and “Out_of_Context” samples across case and control cohorts. Fisher’s exact test demonstrated a statistically significant difference between groups (p = 0.036), with an odds ratio of 0.0 (95% CI: 0.005-1.464). Only 0.68% of H/L cases were BRAF-mutated compared with 7.2% of NHW controls, indicating that early-onset NHW patients treated with FOLFOX were more likely to harbor BRAF mutations than their H/L counterparts. These findings highlight ancestry-associated molecular differences in early-onset CRC and underscore the utility of AI-driven interrogation for uncovering clinically relevant genomic disparities.

**Figure S5.**
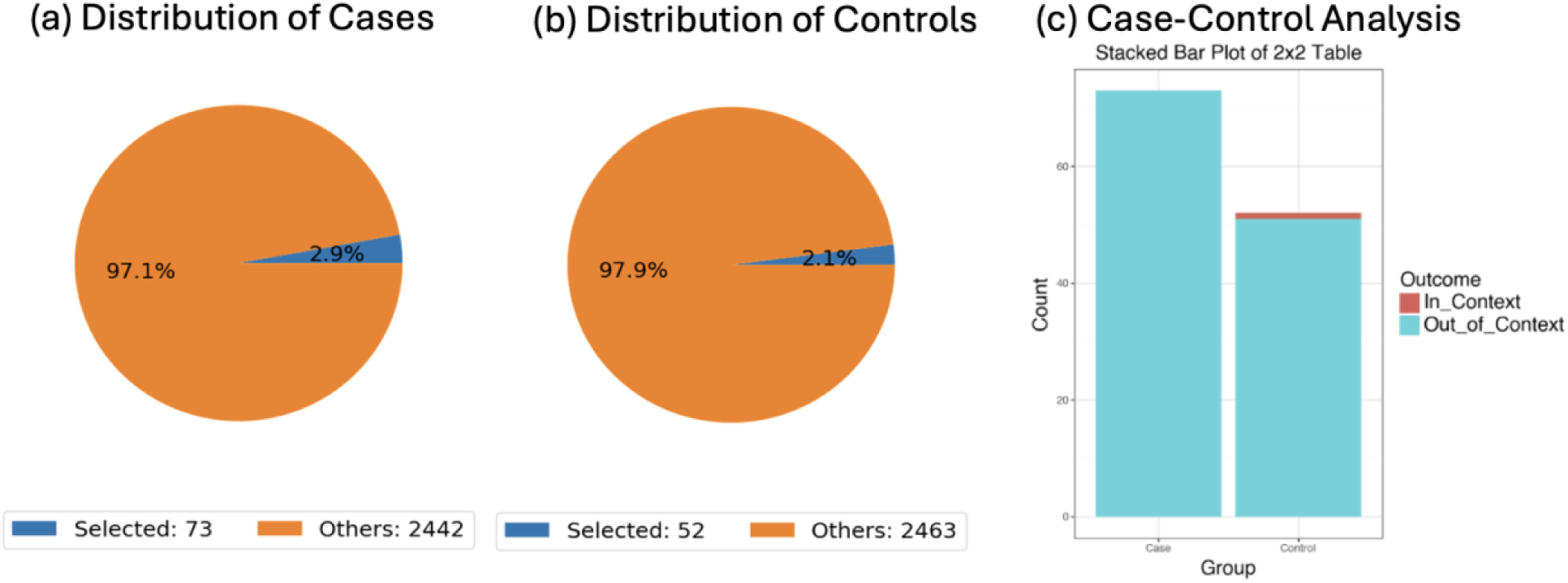
AI-assisted comparison of ERBB2 mutation frequency between early-onset Hispanic/Latino (H/L) colorectal cancer (CRC) patients treated with FOLFOX and early-onset H/L patients not treated with FOLFOX. The AI-HOPE-JAK-STAT platform was used to define case and control cohorts based on user-specified clinical and treatment parameters. The case cohort consisted of early-onset H/L CRC patients who received FOLFOX (n = 73), while the control cohort included early-onset H/L patients who did not receive FOLFOX (n = 52). ERBB2 mutation status was used as the in-context criterion for comparison. The pie charts illustrate the proportion of selected (in-context) versus unselected (out-of-context) samples within each cohort. The stacked bar plot summarizes the number of ERBB2-mutated and non-mutated samples across case and control groups. Fisher’s exact test showed no statistically significant difference in ERBB2 mutation prevalence between treated and untreated early-onset H/L patients (p = 0.864), with an odds ratio of 0.0 (95% CI: 0.012-10.61). These results indicate that ERBB2 mutation frequency does not differ meaningfully by FOLFOX treatment status within early-onset H/L CRC patients.

## Notes

### Competing Interest Statement

The authors have declared no competing interest.

### Funding Statement

This study was funded by the National Cancer Institute, NCI, award number U2CCA252971; the City of Hope Cancer Control and Population Sciences program by the National Institutes of Health, NIH, National Cancer Institute, NCI, award number P30CA033572; and the Drug Development and Capacity Building: A UCR/CoH-CCC Partnership project by the National Institutes of Health, NIH, National Cancer Institute, NCI, award number U54 CA285116.

### Author Declarations

All data used in the present study is publicly available at https://www.cbioportal.org/ and https://genie.cbioportal.org.

